# Cystatin C is glucocorticoid-responsive, directs recruitment of Trem2+ macrophages and predicts failure of cancer immunotherapy

**DOI:** 10.1101/2021.08.17.21261668

**Authors:** Sam O. Kleeman, Breanna Demestichas, Nicholas Mourikis, Dominik Loiero, Miriam Ferrer, Sean Bankier, Yosef J.R.A. Riazat-Kesh, Hassal Lee, Dimitrios Chantzichristos, Claire Regan, Jonathan Preall, Sarthak Sinha, Nicole Rosin, Bryan Yipp, Luiz G.N. de Almeida, Jeff Biernaskie, Antoine Dufour, Pinkus Tober-Lau, Arno Ruusalepp, Johan L. M. Bjorkegren, Markus Ralser, Florian Kurth, Vadim Demechev, Todd Heywood, Qing Gao, Gudmundur Johannsson, Viktor H. Koelzer, Brian R. Walker, Hannah V. Meyer, Tobias Janowitz

**Affiliations:** Cold Spring Harbor Laboratory, Cold Spring Harbor, New York, USA; Department of Pathology and Molecular Pathology, University Hospital Zurich, University of Zurich, Zurich, Switzerland; BHF Centre for Cardiovascular Science, Queen’s Medical Research Institute, University of Edinburgh, Edinburgh, UK; Computational Biology Unit, Department of Informatics, University of Bergen, Bergen, Norway; Mount Sinai Hospital, New York, USA; Department of Internal Medicine and Clinical Nutrition, Institute of Medicine at Sahlgrenska Academy, University of Gothenburg, Gothenburg, Sweden.; Department of Endocrinology Diabetes and Metabolism, Sahlgrenska University Hospital, Gothenburg, Sweden.; Department of Comparative Biology and Experimental Medicine, Faculty of Veterinary Medicine, University of Calgary, Calgary, AB, Canada; Department of Critical Care Medicine, Cumming School of Medicine, University of Calgary, Calgary, AB, Canada; Department of Biochemistry and Molecular Biology and Physiology and Pharmacology, University of Calgary, Calgary, Canada; Department of Comparative Biology and Experimental Medicine, Faculty of Veterinary Medicine and Department of Surgery, Cumming School of Medicine, University of Calgary, Canada; Charité - Universitätsmedizin Berlin, Freie Universität Berlin and Humboldt-Universität zu Berlin, 13353 Berlin, Germany; Department of Cardiac Surgery, Tartu University Hospital, Tartu, Estonia; Department of Genetics & Genomic Sciences, Institute of Genomics and Multiscale Biology, Icahn School of Medicine at Mount Sinai, New York, NY, USA; Department of Oncology and Nuffield Department of Medicine, University of Oxford, Oxford, United Kingdom; Translational & Clinical Research Institute, Newcastle University, Newcastle upon Tyne, UK; Cancer Institute, Northwell Health, New York, USA

## Abstract

Cystatin C (CyC) is a secreted cysteine protease inhibitor and its biological functions remain insufficiently characterized. Plasma CyC is elevated in many patients, especially when receiving glucocorticoid (GC) treatment. Endogenous GCs are essential for life and are appropriately upregulated in response to systemic stress. Here we empirically connect GCs with systemic regulation of CyC. We used genome-wide association and structural equation modeling to determine the genetics of the latent trait CyC production in UK Biobank. CyC production and a polygenic score (PGS) capturing germline predisposition to CyC production predicted elevated all-cause and cancer-specific mortality. We then demonstrated that CyC is a direct target of GC receptor, with GC-responsive CyC secretion exhibited by macrophages and cancer cells. Using isogenic CyC-knockout tumors, we discovered a markedly attenuated tumor growth *in vivo* and found abrogated recruitment of Trem2+ macrophages, which have been previously linked to failure of cancer immunotherapy. Finally, we showed that the CyC-production PGS predicted checkpoint immunotherapy failure in a combined clinical trial cohort of 685 metastatic cancer patients. Taken together, our results demonstrate that CyC may be a direct effector of GC-induced immunosuppression, acting through recruitment of Trem2+ macrophages, and therefore could be a target for combination cancer immunotherapy.

## Introduction

Large prospective patient cohorts with comprehensive genetic, physical and health data, captured in biobanks, allow for the re-evaluation of human disease, health and health care^1^. Due to substantial genetic variation between humans^2^, genome-wide association studies (GWAS) are analogous to forward genetic screens and can direct discovery of the molecular determinants of complex biomedically relevant phenotypes such as organ function^3^. Previously, we have developed a model for the accurate estimation of kidney filtration function, defined as the estimated glomerular filtration rate (eGFR), in patients with cancer^4, 5^. Like others before, we used creatinine^6^, a breakdown product of muscle creatine metabolism that is renally excreted^7^, as a predictor variable. In non-cancer patients, this approach has been compared to the use of cystatin C (CyC; gene name *CST3*)^8^, a secreted cysteine protease inhibitor. Serum levels of both molecules depend on latent (unmeasured) components: most notably their synthesis and externalization by the producing cells and the GFR. While the determinants of creatinine production are relatively well-characterized and relate to muscle mass and diet^9^, the factors that regulate CyC production are in contrast poorly understood^10^. Unlike the metabolic end-product creatinine, but like many other secreted proteins, CyC has biological functions: in its monomeric form, it is a highly potent paracrine inhibitor of cysteine proteases^11, 12^.

Given the known function of CyC and its extracellular localization, it is likely that CyC production is systemically regulated. The first indication of this regulation is from the following set of observations. Organ transplant patients tend to have a higher serum CyC for a given measured GFR^13^. The vast majority of transplant patients are prescribed exogenous glucocorticoids (GCs), such as prednisolone or dexamethasone, as part of their immunosuppressive regimen^14^. Paired analyses accounting for patient-specific factors and renal function have demonstrated that exogenous GCs increase CyC production^15^, an effect that has also been observed in patients with excess endogenous GC production (Cushing’s syndrome)^16^. Moreover, CyC production is increased in a range of disease states that induce GC elevations, including viral infection^17^, inflammatory disease^10^ and cancer^18^. This positive association between GC exposure and CyC production has been recapitulated experimentally *in vitro*^19^ and *in vivo*^20^.

Cortisol, the endogenous GC in humans, is produced by the adrenal gland^21^ in a circadian rhythm peaking in the early morning^22^. Through action on the cytosolic GC receptor (GR), GCs profoundly modulate the cellular transcriptional landscape^23^, affecting up to 20% of all genes^24^ and driving systemic reprogramming of metabolism and immunity that is essential for life^25^. While our understanding of the mechanisms by which GCs are immune-modulatory remains limited^26^, their immunosuppressive function is firmly established and therapeutically employed across a wide range of auto-immune and inflammatory diseases, such as rheumatoid arthritis^27^. They are also used to mitigate immune-mediated damage to normal organ systems, a common and potentially severe side effect of T cell activation by checkpoint immunotherapy (CPI) in cancer^28^. This latter indication has emphasized the importance of determining whether, and in what circumstances, exogenous GCs could impair the efficacy of CPI^29, 30^. Evidence from *in vivo* models of cancer suggests that even low doses of GCs can suppress anti-tumor immunity^30^, leading to enhanced metastasis and reduced survival^31^. This has remained difficult to empirically investigate in cancer patients due to confounding by performance status and comorbidities^32^, inconsistent CPI trial inclusion criteria^29^ and the difficulties in performing well-controlled trials in this context.

We hypothesized that, rather than being a passive marker of renal function, cystatin C is directly associated with disease states and that this association might be mediated by GC signaling. Here, to empirically investigate this question, we leverage UK Biobank (UKB), a prospective population-based cohort comprising approximately 480,000 subjects who provided germline genetics, serum CyC and serum creatinine. Using conventional GWAS for eGFR-CyC/eGFR-creatinine followed by structural equation modeling (SEM) we estimate single nucleotide polymorphism (SNP)-level associations with the latent trait of CyC-production. We characterize patient-level predisposition to CyC-production via construction of a polygenic score (PGS) which is validated in a held-out cohort. Through multi-modal genomics, *in vitro*, *in vivo* and experimental medicine approaches we link CyC to GC signaling, recruitment of Trem2+ macrophages and failure of cancer immunotherapy.

## Results

### Genomic architecture of CyC production

To investigate the genomic architecture of CyC production, we first performed a discovery GWAS for eGFR-CyC and eGFR-Creatinine (eGFR-Cr) in 381,764 European subjects in UKB, using linear mixed models to account for population stratification and cryptic relatedness. We randomly selected 50,000 unrelated subjects from the overall UKB European population and excluded their data from the GWAS to enable later validation analyses (Figure 1b). Using linkage disequilibrium (LD) score regression, we identified a strong genetic correlation (r^2^=0.61) between eGFR-CyC and eGFR-Cr, consistent with both traits sharing a common factor that reflects renal filtration function. We reasoned that the genetic variance in eGFR-CyC that was not explained by this common factor represented the latent trait of CyC-production, given that the CyC plasma level is a function of both CyC excretion in the kidney and its cellular production. Thus, we estimated the single-nucleotide polymorphism (SNP)-level effects on CyC-production and renal function, by constructing a genomic structural equation model (SEM, Figure 1c, Figure S1a-b) implemented in Genomic-SEM^33^, assuming no covariance between CyC-production and renal function. Providing confidence in our approach, loci known to directly regulate renal function such as *SHROOM3*^34^ and *UMOD*^35^ were predominantly associated with the renal function latent trait, while the locus coding for CyC (*CST3*) was predominantly associated with the CyC-production latent trait. Other loci associated with CyC-production, such as *SH2B3*^36^ and *FLT3*^37^, identify components of immune cell signaling cascades and are strongly associated with autoimmune disease. The index SNP at the *SH2B3* locus is a missense variant (R262W) and exhibits a markedly larger effect size than would be expected for its allele frequency (minor allele fraction = 0.48, Figure S1c), consistent with evidence that this variant is under active positive selection^38^. The *CPS1* locus, coding for carbamoyl-phosphate synthase 1, stood out as having divergent effects on renal function and CyC-production, probably reflecting its independent roles in creatine metabolism^39^ and immune signaling^40^. We next performed tissue-specific partitioned heritability analysis using gene expression and chromatin accessibility datasets (including GTEx^41^ and Roadmap Epigenomics Project^42^) and this confirmed enrichment of heritability of the renal function rather than CyC- production component of CyC levels in kidney tissues (Figure S1d-e). This analysis also demonstrated enriched heritability for the renal function trait in liver tissues, which may reflect the coupling of hepatic and kidney function, observed clinically as hepatorenal syndrome^43^.

**Figure 1.**
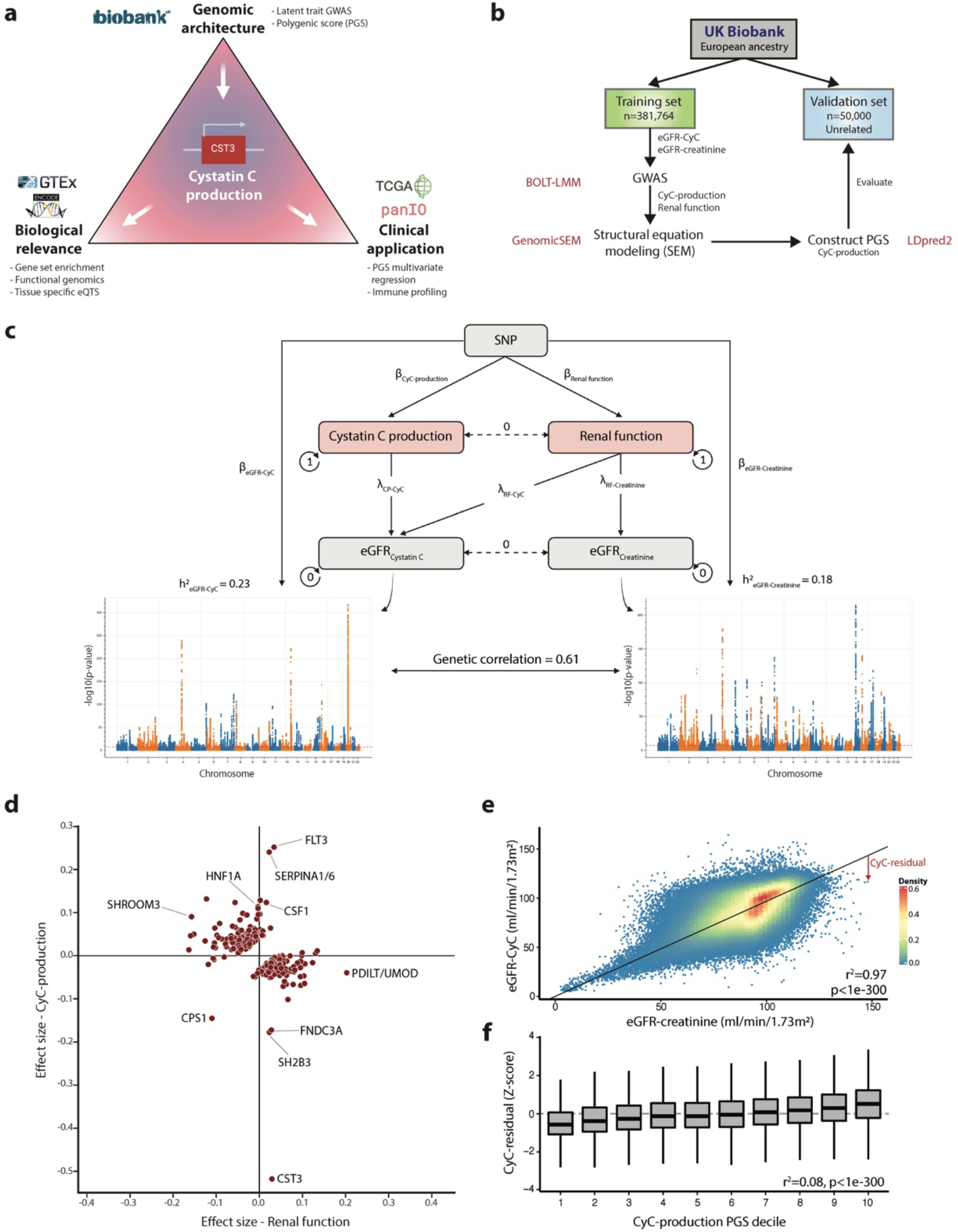
Genomic architecture of cystatin C (CyC) production. (a) Schematic of study plan. The analysis of CyC-production latent trait in UK Biobank (UKB) is leveraged to determine the biological and clinical relevance of CyC. (b) Consort diagram and summary of UKB genome-wide association analysis strategy in the European ancestry population. The software packages utilized for each step are displayed in red. (c) Structural equation model to estimate latent traits of CyC-production and renal function. The model schematic, heritability (h^2^) of eGFR-creatinine and eGFR-CyC, and their genetic correlation derived from LD score regression are shown. Circular arrows refer to variance of each component and dashed lines refer to covariance between components. RF, Renal Function. (d) Latent trait effect sizes (CyC-production and renal function) for single nucleotide polymorphisms (SNPs) corresponding to each clumped locus in eGFR-CyC summary. Gene names are annotated per OpenTargets V2G pipeline. (e) Linear model of eGFR- CyC as a function of eGFR-creatinine across all paired blood samples in UK Biobank, including sex as a covariate. The deviation of the eGFR-CyC from the linear fit as indicated by the red arrow is defined as the CyC-residual, a surrogate for CyC production. (f) Correlation of CyC-residual with CyC-production polygenic score (PGS). Only data from the independent validation set (see panel b) were used. Boxplots show median (central line) with interquartile range (IQR, box) and extrema (whiskers at 1.5× IQR).

Using the discovery data set, we captured the polygenic architecture of CyC-production by deriving a polygenic score (PGS), implemented in LDpred2^44^ using HapMap3 variants, that could be reliably imputed in all of UKB, The Cancer Genome Atlas (TCGA) and Genotype-Tissue Expression (GTEx) cohorts (Figure S1f, Supplemental Data). We sought to maximize portability to clinical sequencing cohorts where only exome sequencing is available, and thus derived a second PGS from HapMap3 variants that could be reliably imputed from exome sequencing data (Methods, Figure S1g). To validate both PGSs with the data from the 50,000 unrelated European patients (Figure 1b), it was necessary to define an independent patient-level estimate for CyC- production. This is possible because the discordance between eGFR-CyC and eGFR-Cr can be viewed as an approximation for CyC-production. Therefore, we modelled eGFR-CyC as a function of eGFR-Cr and sex, and computed the residual (termed CyC-residual, Figure 1e). Using this CyC- residual as a surrogate for CyC-production, we confirmed that the genome-wide CyC-production PGS had significant predictive power in the validation cohort (r^2^=0.08, p<1e-300, Figure 1f). As expected, predictive performance was reduced for the exome-wide PGS in the validation cohort (r^2^=0.04, p<1e-300).

To investigate the trans-ancestral portability of the genome-wide CyC-production PGS, we measured performance versus CyC-residual in African (AFR, n=8152) and Central and South Asian (CSA, n=9845) genetic ancestry groups in UKB. We observed poor trans-ancestral portability of this PGS in these ancestry groups, with a low proportion of CyC-residual variance explained in CSA and AFR populations (Figure S2a-b). In order to derive a PGS in each non-EUR population, we performed GWAS and SEM as described above (Figure 1b) in these two ancestry groups but these analyses were underpowered to detect any signals reaching genome-wide significance (Figure S2c-d). While the genetic correlation between eGFR-CyC and eGFR-Cr in CSA subjects (r^2^=0.65) was comparable to EUR subjects (r^2^=0.61), genetic correlation was substantially diminished in AFR subjects (r^2^=0.18). This indicates that eGFR-Cr and/or eGFR-Cy correlate weakly with true GFR in the AFR population, thus providing empirical genetic evidence to the observation that eGFR models have reduced performance in individuals self-identifying as Black or African American^45^.

### CyC production is associated with accelerated onset of disease

We hypothesized that these quantitative measures of CyC production (CyC-residual and CyC- production PGS) could be used to empirically investigate its prognostic potential. As a preliminary analysis, we used multivariate Cox regression to estimate the effect of CyC-residual on all-cause mortality, adjusted for relevant patient covariates known to predict mortality^46–48^. We found that CyC-residual was associated with significantly increased all-cause mortality (HR=1.56, p<1e-16, Figure 2a). We considered that CyC-residual has the potential to be confounded by environmental factors, including exogenous GC treatment and, to mitigate this, we investigated whether the germline predisposition to CyC production, estimated as CyC-production PGS, could predict lifespan in our UKB European validation set (Figure 1b). Using multivariate Cox regression adjusted for sex, year of birth and principal components capturing genetic ancestry, we found that CyC-production PGS was associated with significantly reduced lifespan of UKB subjects (p=0.00013), as well as their two parents (p<1e-16, Figure 2b).

**Figure 2.**
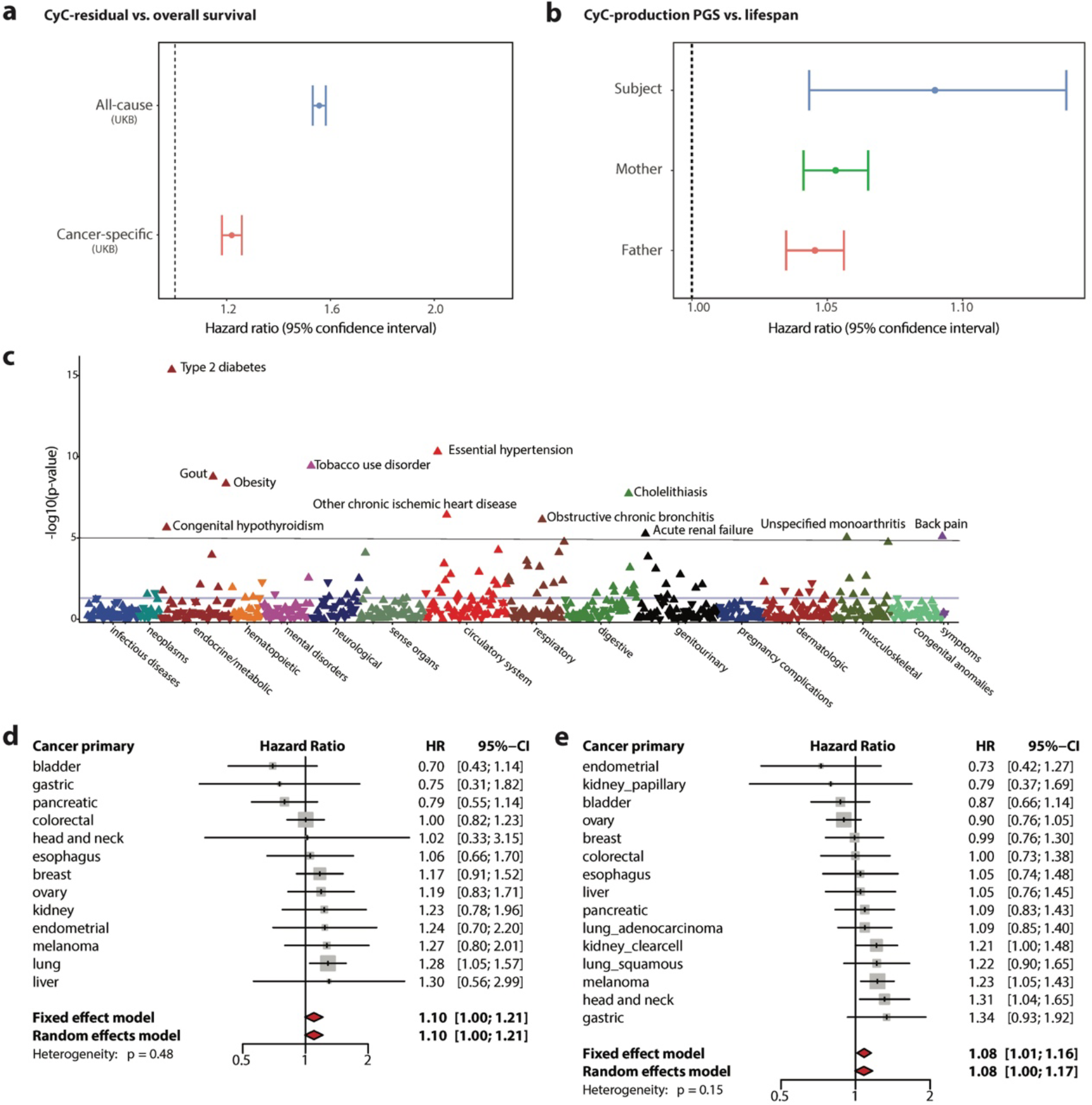
Cystatin C (CyC) production is associated with multiple disease states and is prognostic in cancer patients. (a) Multivariate Cox regression to measure effect size for CyC- residual on overall survival in UK Biobank. Covariates included age, sex, body mass index (BMI), hemoglobin, C-reactive protein, eGFR-creatinine and operation status (for cancer-specific sub-analysis). (b) Multivariate Cox regression to measure effect size for CyC-production polygenic score (PGS) on subject and parental lifespan in UKB validation cohort. Covariates included PC1-4, recruitment center, genotyping array, year of birth of subject and sex of subject (if applicable). (c) Phenome-wide association (Cox regression) between CyC-production PGS and 694 time-to-event phenotypes in UKB validation cohort. Covariates included PC1-4, year of birth and sex. Multivariate Cox regression to measure effect size for CyC-production PGS on disease-specific survival for specific cancers in (d) UKB validation cohort (cancers diagnosed since 2000, n=3954) and (e) TCGA cohort (n=4368). Covariates included PC1-4, age, sex and a term reflecting whether patient had curative surgery. Error bars indicate 95% confidence interval; grey squares indicate sample size.

We considered that increased all-cause mortality might be explained by either earlier onset of specific disease states, or reduced prognosis following disease diagnosis. To investigate the former, we performed a phenome-wide association analysis (PheWAS) in the UKB validation set to identify time-to-event phenotypes (n=694) that were significantly associated with CyC- production using multivariate Cox regression (Figure 2c). We identified positive associations meeting phenome-wide significance (p<1e-5) between CyC-production PGS and multiple diseases linked to metabolic syndrome, including type 2 diabetes, obesity, hypertension and ischemic heart disease. To investigate how CyC-production could modulate disease prognosis, and as elevated plasma CyC is associated with cancer^18^, we examined whether CyC-residual and CyC-production PGS were independent predictors of adverse outcomes in patients with cancer. Using UKB patients diagnosed with cancer since 2000 and with cancer-specific mortality defined by manual review of death certificates, we found that CyC-residual is an independent predictor of increased cancer-specific mortality (HR = 1.22, p<1e-16, Figure 2a). To validate this finding, we performed multivariate Cox regression of cancer-specific mortality against CyC-production across 13 tumor groups in 2 independent cohorts (UKB validation set, TCGA European subjects). Both fixed and random effect meta-analyses in each independent cohort confirmed a significant positive association between CyC-production PGS and cancer-specific mortality (Figure 2d-e). We noted that while there was variation in a single-cancer level the overall effect size was concordant between UKB and TCGA. Consistent with this, we have found that CyC-production PGS is associated with increased odds of COVID-19 critical illness in four cohorts spanning European and African ancestry populations (manuscript under review at *iScience* and attached to this submission). In summary, the association between CyC-production PGS and reduced lifespan is likely to be explained by a combination of earlier disease onset and reduced disease-specific survival.

### CyC is a glucocorticoid response gene *in vitro*

To better understand the mechanism by which CyC production could regulate disease incidence and prognosis, we reviewed the genetic loci most associated with CyC-production in our GWAS summary statistics. The *SERPINA1/6* locus on chromosome 14 had one of the largest effect sizes for CyC-production (Figure 1d, Figure 3a) and is known to be associated with plasma cortisol^49^, implying the possibility of a link between cortisol and CyC. In a recent cortisol Genome-Wide Meta-Analysis (GWAMA), this signal was thought to be mediated by altered hepatic expression of *SERPINA6*^49^, which encodes cortisol-binding globulin (CBG). To determine if there was a shared common variant, we performed co-localization analysis^50^. We did not detect a shared causal variant (posterior probability = 1.45e-15), but trans-expression qualitative trait loci (trans-eQTL) analysis in the Stockholm Tartu Atherosclerosis Reverse Networks Engineering Task (STARNET)^51^ cohort identified a single SNP (rs2749527) at the *SERPINA1/6* locus that was associated with significantly reduced plasma cortisol (p=1.75e-13) and significantly reduced *CST3* gene expression in visceral adipose fat (p = 0.0024 in additive model, p=9.21e-6 in recessive model, Bonferroni-adjusted alpha level of 0.0025, Figure 3b). This variant is independently associated with significantly reduced liver *SERPINA6* expression in STARNET (p=4.73e-9, Figure 3c) and GTEx (p=0.004, Figure S3c) cohorts. As such, a single genetic instrument connects CBG, plasma cortisol and CyC (Figure S3a), thus providing genetic evidence for a direct link between GCs and CyC.

**Figure 3.**
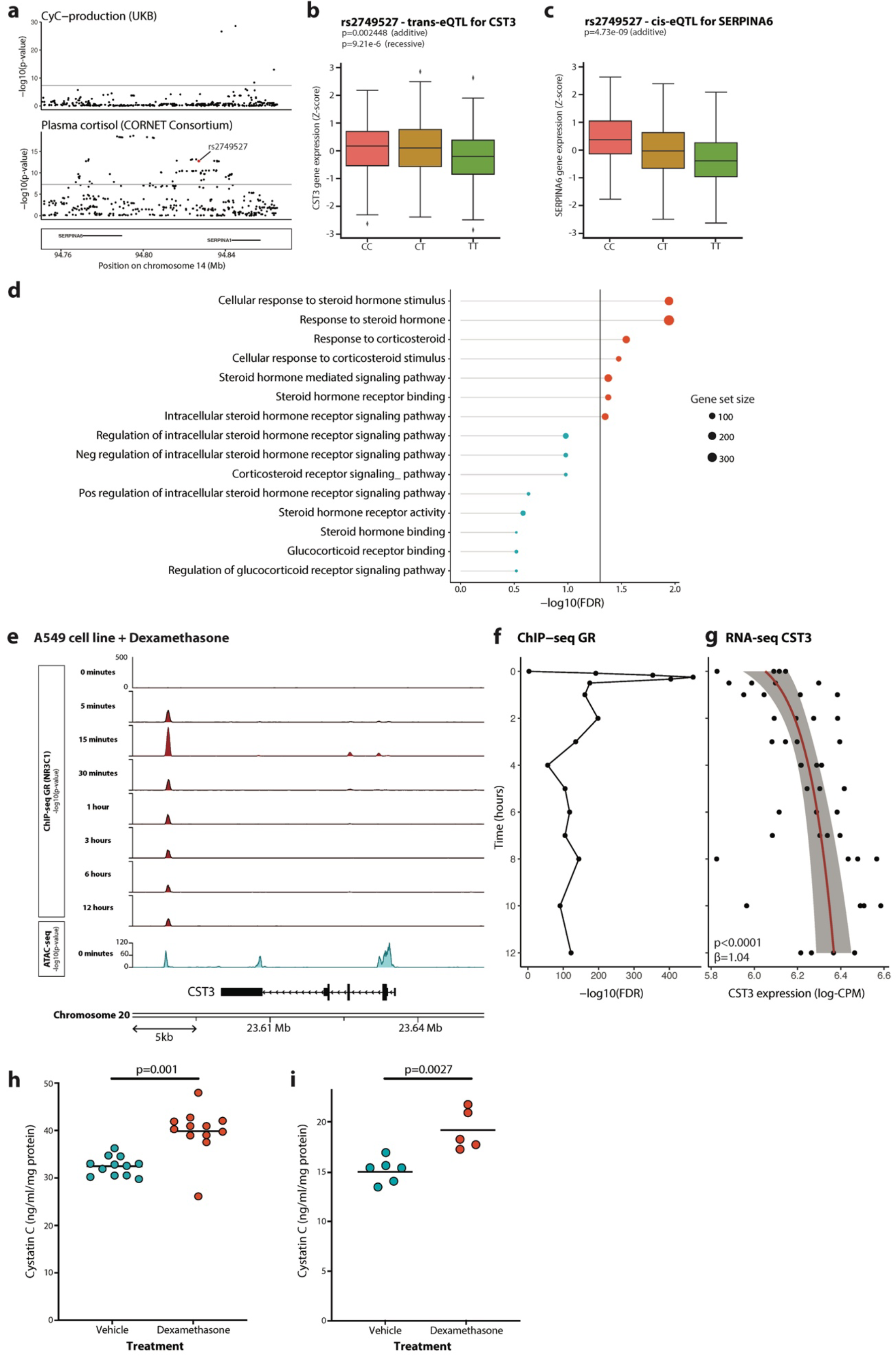
Cystatin C (CyC) is a glucocorticoid response gene *in vitro*. (a) Colocalization of summary statistics for CyC-production from UK Biobank (UKB) and plasma cortisol from CORNET Consortium at *SERPINA1/6* locus. rs2749527 variant is highlighted in red. (b) Trans-eQTL analysis examining association between genetic instrument rs2749527 and *CST3* gene expression in visceral adipose fat (VAF) in STARNET cohort. (c) Cis-eQTL association between rs2749527 and *SERPINA6* (encodes cortisol-binding globulin) in liver in STARNET cohort. See Figure S3 for replication analysis in GTEX. (d) Gene set enrichment analysis (MAGMA) across CyC-production summary statistics (UKB) for steroid signaling-related gene sets. (e) Functional genomics in A549 cell line (ENCODE project) treated with 100nM dexamethasone for 0 minutes to 12 hours. CHIP- seq (for glucocorticoid receptor/NR3C1) and ATAC-seq (at 0 hours) at *CST3* locus identifies a glucocorticoid-responsive and accessible distal enhancer element. Timecourse of (f) GR recruitment (at distal enhancer) and (g) *CST3* gene expression (log-CPM) following dexamethasone treatment in A549 cells (ENCODE project). Trendline and shaded 95% confidence interval correspond to regression of gene expression as a function of log-time. Extracellular cystatin C concentration in (h) A549 cells and (i) HeLa cells normalized to cellular protein content after 18-hour treatment with 100nM dexamethasone or vehicle control. Each condition comprises at least 5 biological replicates; horizontal bars indicate mean extracellular CyC. See Figure S3 for timecourse. (b, c) Boxplots show median (central line) with interquartile range (IQR, box) and extrema (whiskers at 1.5× IQR). Outliers beyond 1.5× IQR are shown as dots.

To further examine the link between GCs and CyC, we mapped each SNP meeting genome-wide significance to overlapping genes (defined by transcriptional start and end sites) and performed gene set enrichment analysis (GSEA) for gene sets relating to GC signaling. This analysis identified significant enrichment of 7/15 GC signaling gene sets from the Gene Ontology Resource (Figure 3d). In light of this, we hypothesized that *CST3* might be a direct transcriptional target of glucocorticoid receptor (GR, gene name *NR3C1*). Using functional genomics data derived from the ENCODE project, including chromatin immunoprecipitation sequencing (ChIP-seq) for GR and Assay for Transposase-Accessible Chromatin using sequencing (ATAC-seq) data in the A549 cell line treated with dexamethasone, we identified dexamethasone-induced recruitment of GR to an accessible downstream enhancer element at the *CST3* locus (Figure 3e-f). In the same experiment, dexamethasone significantly increased *CST3* gene expression over time (p<0.0001, Figure 3g). We sought to investigate whether, and on what timescale, the transcriptional induction of *CST3* by dexamethasone results in increased cellular secretion of CyC, which would cause increased tissue and circulating CyC levels. We first repeated the ENCODE experimental protocol using A549 cells and found that extracellular CyC concentration was significantly increased after 18 hours of treatment compared to 0 hours (Figure S3d). We next compared extracellular CyC concentration 18 hours after treatment with either dexamethasone or vehicle control in A549 cells (Figure 3h) and HeLa cells (Figure 3i) and found significant elevations of CyC in both cases.

In light of our finding that extracellular CyC concentrations do not significantly increase until 18 hours after dexamethasone treatment *in vitro* (Figure S3d), we hypothesized that CyC might be able to smooth the diurnal rhythm in plasma cortisol levels. Despite the fact that plasma cortisol was not directly measured in UKB, we were able to assess this, because previous studies have demonstrated that plasma bilirubin is strongly correlated to plasma cortisol over time^52^. Using cosinor regression^53^ for CyC-residual and bilirubin in UKB subjects with available blood sampling, we determined that the diurnal variation of CyC-residual is diminished compared to bilirubin (amplitude coefficient = 0.038 versus 0.23, Figure S3e). Moreover, the peak in CyC-residual appears to be delayed by approximately 18 hours compared to the expected peak in plasma cortisol, consistent with our *in vitro* experiments. Taken together, these findings demonstrate that CyC production is directly induced by GCs, and that CyC has a reduced diurnal amplitude and offset periodicity compared to plasma cortisol.

### CyC is secreted in healthy individuals by monocytes in a glucocorticoid independent manner

CyC has been validated as an effective marker of renal function in multiple large clinical cohorts^54^ comprising patients without acute disease but has been shown to be dynamically regulated in disease states such as cancer^18^, and so we hypothesized that GC-inducible expression of CyC would operate in a context-dependent manner. To investigate this hypothesis, it was first necessary to characterize the dominant source of secreted CyC in health. At first glance, *CST3* gene expression was relatively consistent across all tissues examined as part of the GTEX project (GTEX Portal), but we reasoned that tissues that predominantly secrete CyC would exhibit a significant positive correlation between CyC-production PGS and *CST3* gene expression. Using expression qualitative trait score (eQTS) analysis we detected a significant positive correlation in spleen tissues (n=171, Figure 4a). In support of this, we identified circadian rhythmicity from cosinor regression of spleen *CST3* gene expression against time of death, which was attenuated compared to the canonical GC target *FKBP5* (amplitude = 0.060 versus 0.24, Figure S4a). To understand which cell types might be driving this signal, we examined available single-cell RNA sequencing (scRNA-seq) data from human spleen^55^. This showed that only myeloid-derived cell populations (dendritic cells, macrophages and monocytes) expressed *CST3* (Figure 4b). We confirmed myeloid-specific *CST3* expression in peripheral blood mononuclear cells (PBMCs) scRNA-seq^56^ (Figure 4c) and across multiple scRNA-seq datasets harmonized as part of the Human Protein Atlas^57^ (Figure 4d). As additional validation supporting the role of monocytes as a dominant contributor to plasma CyC levels, we found a significant positive correlation between blood monocyte counts and CyC-residual in the UKB cohort (Figure S4b), while two-sample Mendelian randomization using blood-derived *CST3* eQTLs (eQTLGen^58^) as exposure identified a highly significant positive association with CyC-production (p=6.13e-77, Figure 4e). On the basis of these results, we hypothesized that monocyte-like THP-1 cells would have high basal *CST3* gene expression and secretion of CyC without GC-inducibility, and we confirmed this by RT-PCR (Figure 4f) and ELISA (Figure 4g). Consistent with these findings, dexamethasone treatment did not elevate plasma CyC levels in healthy BALB/c (Figure 4h) and C57BL/6J (Figure 4i) mice, nor did near-physiological hydrocortisone treatment affect creatinine-normalized CyC levels (ratio of eGFR-Creatinine to eGFR-CyC, termed C2 ratio) in patients with primary adrenal insufficiency^59^ (Figure 4j). Altogether, these findings indicate that CyC production is relatively constant in health, and in health does not significantly increase in response to GC agonism, explaining the validated utility of CyC as a marker of renal function in patients without acute illness.

**Figure 4.**
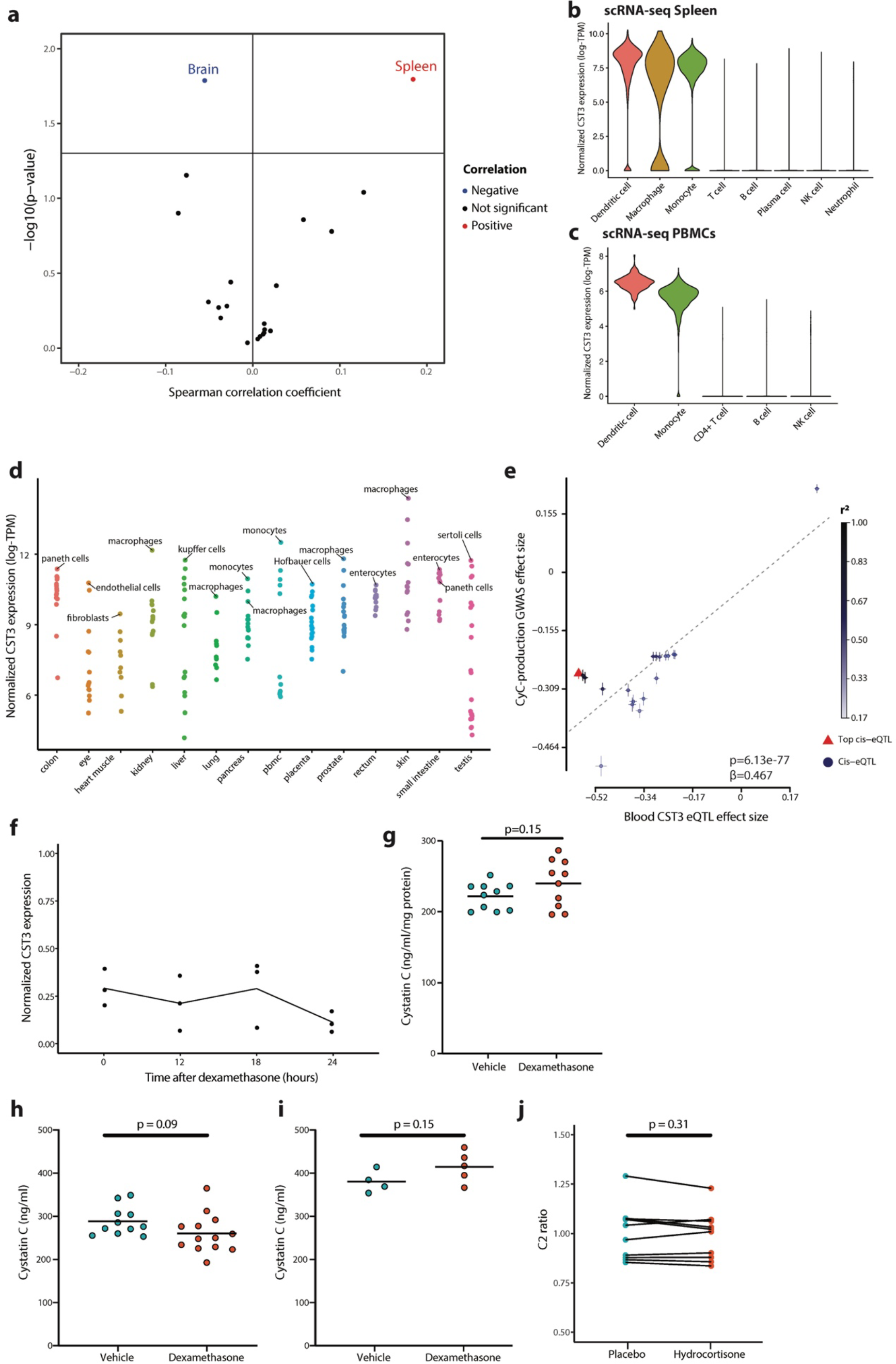
Cystatin C (CyC) is predominantly produced by myeloid cells in health. (a) Tissue-specific expression quantitative trait score (eQTS) analysis to identify tissues with significant correlation (Spearman coefficient) between CyC-production polygenic score (PGS) and tissue-specific *CST3* gene expression in GTEX cohort. P values are uncorrected as each correlation test is performed in a non-overlapping set of tissue-specific samples. Distribution of normalized single-cell *CST3* expression (log-transcripts per million [TPM]) in cell clusters isolated from (b) spleen and (c) peripheral blood mononuclear cells (PBMCs). Clusters defined by correlation to reference PBMC data^91^. (d) Mean *CST3* gene expression (log-TPM) in each cell cluster from multiple tissue-specific single-cell RNA sequencing projects, harmonized by Human Protein Atlas. The top cell cluster and tissue-specific macrophage cell type (if not top cluster) by tissue is annotated. (e) Two-sample mendelian randomization using blood-specific cis-eQTLs for *CST3* (eQTLGen) as exposure and CyC-production latent trait GWAS as outcome. Error bars correspond to standard errors, point color refers to linkage with top cis-eQTL. Change in (f) *CST3* gene expression (reverse transcription-PCR) and (g) extracellular CyC concentration in human THP-1 cells (monocyte-like) normalized to cellular protein content after 18-hour treatment with dexamethasone (100 nM) or vehicle control. Each condition comprises 10 biological replicates. Plasma CyC concentration in healthy (h) BALB/cJ and (i) C57BL/6J mice treated with vehicle or 20mg/kg dexamethasone. (j) Creatinine-normalized plasma CyC (C2 ratio) in patients with primary adrenal insufficiency treated with placebo (saline) or hydrocortisone in a crossover experimental medicine study. The administered IV hydrocortisone dose was 0.030 mg/kg/hr between 12 AM and 7 AM (the time point of sampling), achieving near-physiological GC exposure.

### CyC secretion is dynamically and glucocorticoid dependently regulated in disease states

Inflammation is characterized by the recruitment of monocytes to diseased tissues, where they differentiate into macrophages^60^. As GR is expressed in macrophages but not in monocytes^61^, we hypothesized that, while monocytes have high constitutive basal CyC production, macrophages would secrete CyC in response to GC signaling. To investigate this question, we treated monocytic human THP-1 cells with the protein kinase C activator PMA (phorbol 12-myristate 13-acetate) to induce macrophage-like differentiation. Dexamethasone treatment of PMA-activated THP-1 cells significantly increased *CST3* gene expression at 12 hours (Figure 5a) and extracellular CyC protein concentration at 18 hours (Figure 5b), mirroring the results found in A549 and HeLa cells (Figure 3h-i). Severe COVID-19 infection is characterized by persistent lung inflammation associated with concomitant recruitment of monocyte-derived macrophages^62^. Until the release of the RECOVERY trial^63^, patients with severe COVID-19 were not routinely treated with GC agonists such as dexamethasone. As such, COVID-19 presents a unique opportunity to investigate the effect of dexamethasone on creatinine-normalized CyC levels (C2 ratio). We collated plasma creatinine and CyC measurements in two independent cohorts of patients (from Calgary, Canada^64^ and Berlin, Germany^65^). In each cohort, a subset of patients received standard of care (pre-RECOVERY trial) and a subset received standard of care plus dexamethasone from admission (post-RECOVERY trial). We identified significantly increased C2 ratios in dexamethasone-treated patients at early timepoints (day 1 or day 3, Figure 5c-d) that normalized by day 7 after admission.

**Figure 5.**
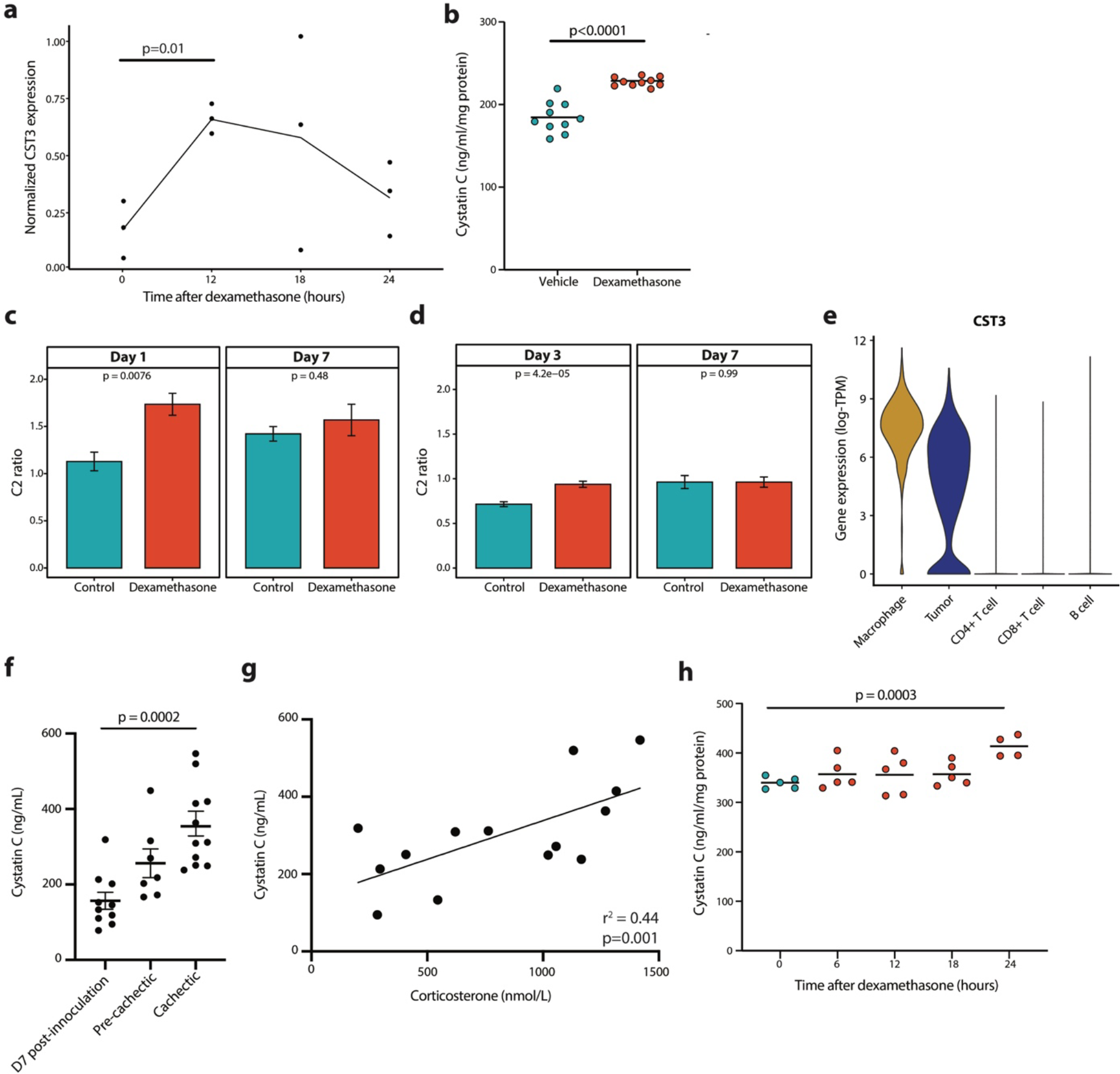
Cystatin C (CyC) production is dynamically regulated in disease states. Change in (a) *CST3* gene expression (reverse transcription-PCR) and (b) extracellular CyC concentration in PMA- treated human THP-1 cells (macrophage-like) normalized to cellular protein content after 18- hour treatment with dexamethasone (100 nM) or vehicle control. Creatinine-normalized plasma CyC (C2 ratio) at specific timepoints with sufficient data in hospitalized COVID-19 patients treated with dexamethasone or standard of care as part of cohorts based in (c) Calgary, Canada and (d) Charité Hospital, Germany. Day 1 in the Calgary, Canada cohort refers to a time window of 72 hours after admission to ICU. (e) Single-cell *CST3* gene expression in each cell cluster in melanoma tumors (n=12) from Jerby-Anon et al^92^. Clusters defined by correlation to reference PBMC data^91^, with unclassified cells that exhibit detectable clonal copy number variation classified as tumor. (f) Plasma CyC concentration in BALBc mice after inoculation with colon-26 (C26) tumor cells. Cachexia defined by >15% body weight loss, pre-cachexia refers to 14 days after tumor inoculation. (g) Significant positive correlation between plasma corticosterone and plasma cystatin C during tumor progression in C26 model. (h) Extracellular cystatin C concentrations in C26 cells normalized to cellular protein content after 0-, 6-, 12-, 18- or 24-hour treatment with 100 nM dexamethasone. Each timepoint comprises at least 4 biological replicates.

The findings that CyC is primarily expressed by myeloid cells and that GC-responsive CyC secretion is predominantly mediated by macrophages have the potential to explain our finding that rs2749527 is a trans-eQTL for *CST3* measured in visceral adipose fat (VAF) in the STARNET but not GTEX (p=0.77, Figure S3b). The STARNET study recruited patients with established coronary artery disease^51^, while GTEX is a relatively unselected cohort of deceased donors^41^. As metabolic syndrome is associated with significant macrophage accumulation in adipose tissue^66^, we hypothesized that STARNET patients would have significantly increased macrophage gene signatures in VAF, compared to GTEX donors. Using CIBERSORTx^67^ (absolute mode) analysis of RNA-seq data in each cohort, we identified highly significant enrichment of M2-like macrophages (demarcated by high expression of *CCL18, TREM2 and CLEC4A)* in STARNET versus GTEX (p=3.03e- 289, Figure S5a). M2-like macrophages were by far the most abundant myeloid component in the STARNET VAF samples, suggesting that they are the cell type underlying the trans-eQTL signal. This finding both provides orthogonal validation for the role of macrophages in GC-responsive CyC secretion and illustrates the limitations of eQTL analysis using bulk RNA-seq data, as has been described previously^68^.

While we did not identify significant *CST3* gene expression in epithelial tissues in the GTEX and Human Protein Atlas datasets, it is notable that we detect high and GC-inducible CyC expression in cancer cell lines (Figure 3h-i). This raises the possibility that cancer cells co-opt a phenotype normally exhibited by macrophages and ectopically express *CST3*. In support of this, we re- analyzed scRNA-seq data from melanoma specimens from 12 patients, confirming high *CST3* expression in the myeloid compartment and identifying comparable ectopic *CST3* expression in the tumor compartment (Figure 4h). Consistent with elevated intratumorally GC levels^26^, Expression of the canonical GC target *FKBP5* could be identified in all cell populations profiled in the tumor (Figure S5b)., demonstrating the GC signaling is necessary but not sufficient for *CST3* expression. The murine colorectal-26 (C26) model of cancer progression is characterized by marked elevations in endogenous GC production during disease progression^30^. As such, we hypothesized and subsequently confirmed that CyC levels would significantly increase during disease progression (Figure 5f), and that these increases would positively correlate with levels of the endogenous murine GC, corticosterone (Figure 5g). Dexamethasone treatment of C26 cells *in vitro* was associated with significantly increased CyC secretion at 24 hours (Figure 5h), suggesting that elevations in CyC during C26 cancer progression are mediated by GC-induced cancer cell-intrinsic CyC secretion. Altogether these findings demonstrate that the capacity of GCs to induce CyC secretion is highly context-dependent, and can be co-opted by cancer cells, suggesting a possible selective advantage to cancer cells.

### CyC directs recruitment of Trem2+ macrophages and failure of cancer immunotherapy

To investigate how CyC expression would provide a selective advantage to cancer cells, we used transient transfection with Cas9 and *CST3*-specific guide RNAs (gRNAs) to generate a CST3- knockout (*CST3^-/-^)* clone of the Mm1 cell line, which is derived from a liver metastasis of the autochthonous KPC model of pancreatic cancer^69^. Knockout was confirmed by ELISA for extracellular CyC (Figure S6a) and Sanger sequencing of the predicted edit site (Figure S6b), which confirmed 97% editing efficiency. While isogenic sgScrambled and *CST3^-/-^* Mm1 clones had equivalent doubling times *in vitro* (sgScrambled: 23.3 hours, 95% CI 21.6-25.4; *CST3^-/-^*): 24.0 hours, 95% CI 22.4-25.8, Figure S6c), *CST3^-/-^* tumors had markedly attenuated growth kinetics *in vivo* (Figure 6a, independent replication Figure S6d) and significantly lower endpoint tumor weights (Figure S6e, independent replication Figure S6f). Considering that this growth defect was only detectable *in vivo*, CyC’s known function as a potent inhibitor of cysteine proteases^12^, such as those involved in antigen presentation^70^, and the role of GCs on CyC secretion, we hypothesized that CyC might have an immune suppressive function. To minimize the effect of mouse-specific factors and to maximize the immune selective pressure on tumors, we inoculated mice with a sgScrambled tumor on the left flank and a *CST3^-/-^* tumor on the right flank (termed bi-flank model) and we treated mice with 2-3 doses of anti-PD-L1 antibody. This experiment confirmed the suppressed growth of *CST3^-/-^* versus paired sgScrambled tumors (Figure 6b and Figure S6h, independent replication Figure S6g and S6i). Consistent with this, the proportion of Ki67-positive cells was significantly lower in *CST3^-/-^* versus paired sgScrambled tumors (Figure 6c).

**Figure 6.**
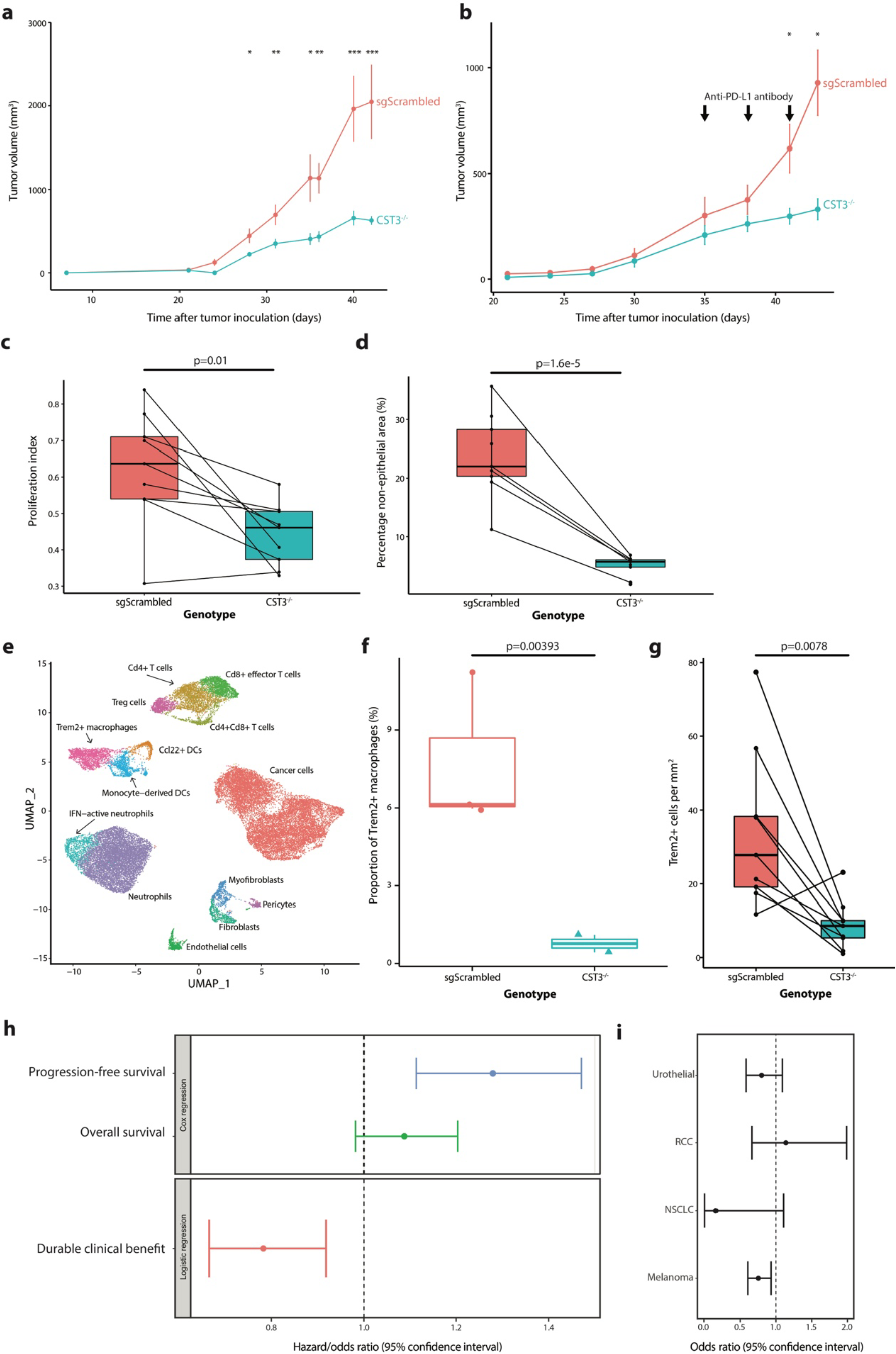
Cystatin C directs recruitment of TREM2+ macrophages and promotes failure of cancer immunotherapy. (a) Tumor growth curves (mean and standard error of the mean) for single-flank sgScrambled (n=8) and *CST3^-/-^* (CST3-KO, n=8) tumors, 100,000 cells inoculated in right flank (cohort A). (b) Tumor growth curves (mean and standard error of the mean) for bi- flank paired sgScrambled (n=5) and *CST3^-/-^* (n=5) tumors, 50,000 cells inoculated in both flanks (cohort C). Mice received three doses of anti-PD-L1 antibody. (c) Proliferation index (proportion of Ki67-positive cells / total cells) and (d) proportion of non-epithelial cells in histological sections from paired bi-flank sgScrambled and *CST3^-/-^* tumors (pooled cohorts C and D). (e) UMAP projection of 14,416 cells, annotated with cell type, from 4 tumor samples (2 sgScrambled, 2 *CST3^-/-^*). (f) Proportion of Trem2+ macrophages in sgScrambled and *CST3^-/-^* tumors, p-value is adjusted p-value from linear model of logit-transformed proportions. (g) Number of Trem2+ cells per mm^2^ from digital image analysis of Trem2 immunohistochemistry in paired bi-flank sections from sgScrambled and *CST3^-/-^* tumors. (h) Multivariate (Cox and logistic) regression of Z-scored CyC-production polygenic score (PGS) against immuno-oncology biomarkers (progression-free survival, overall survival, durable clinical benefit) in meta-analysis of European patients (n=685) treated with checkpoint immunotherapy (anti-CTLA4 or anti-PD1/PD-L1). Sample sizes for each clinical end-point were n=342, n=685 and n=670 respectively. In each model, covariates included the first four principal components, sex and cancer primary. Higher hazard ratios (survival) or lower odds ratios (durable clinical benefit) reflect worse therapeutic outcomes. (i) Sensitivity analysis indicating odds ratio and confidence interval for durable clinical benefit in each cancer type. *, p<0.05; **, p<0.01, ***, p<0.001.

To investigate whether altered growth kinetics reflected remodeling of the tumor microenvironment, we performed pan- cytokeratin immunohistochemistry and automated image segmentation to score the epithelial and non-epithelial areas in each tumor section. The fraction of non-epithelial cells was markedly reduced in paired *CST3^-/-^* versus sgScrambled tumors. In order to identify whether the depletion of specific non-epithelial cell types could explain this observation, we perform single-cell RNA sequencing (scRNA-seq) on 2 sgScrambled and 2 *CST3^-/-^* uni-flank tumors, with 14,416 cells spanning 14 cell types passing quality control criteria (Figure 6e, marker genes used to define cell identity summarized in Figure S7a and Table S7). scRNA-seq profiles of cancer cells confirmed *Cst3* knockout in this compartment (Figure S7b, p=9.16e-30), but not in other compartments (p>0.05). To identify enriched or depleted cell types, we implemented the *propeller* method^71^ which models the logit-transformed cell type proportions as a function of the *CST3* genotype. At 5% FDR we identified a single cell population, annotated as Trem2+ macrophages, which was significantly depleted in *CST3^-/-^* tumors (adjusted p=0.004, ratio=0.098, Figure 6f). We orthogonally validated depletion of Trem2+ cells by digital image analysis of Trem2 immunohistochemistry (IHC) in a non-overlapping cohort of bi-flank sgScrambled and *CST3^-/-^*tumor sections (Figure 6g). Unexpectedly, we identified a highly non-random distribution of Trem2+ cells in both sgScrambled and *CST3^-/-^* sections, with a marked enrichment of Trem2+ cells in the outer rim of the tumor (Figure S7b). These findings suggest that CyC can influence migration or expansion of Trem2+ macrophages, and that Trem2+ macrophages might regulate trafficking of immune cells into the tumor.

As Trem2+ monocytes can be detected in blood samples^72^, we hypothesized that GC treatment in unwell patients would be associated with expansion of Trem2+ monocytes in blood. To investigate this, we re-analyzed CD14+ monocyte scRNA-seq profiles from ICU-admitted patients with COVID-19^64^, recovered dropped-out features using an established approach^73^ and identified a high-confidence Trem2+ monocyte cluster (cluster 0, Figure S7d). We identified significant expansion of cluster 0 at day 7 versus day 1 in patients treated with dexamethasone (adjusted p=0.004, ratio=16.1, Figure S7e). This would support a step-wise model in which GC agonists increase extracellular CyC levels (Figure 5c), which in turn promote recruitment or expansion of Trem2+ myeloid cells.

Others have shown that TREM2+ macrophages play a highly immunosuppressive role in the tumor microenvironment^74^ and are known to be associated with failure of CPI targeting the PD- 1/PD-L1 axis^75, 76^. In light of this, we hypothesized that increased CyC, by inducing recruitment and/or expansion of Trem2+ macrophages, would be associated with reduced efficacy of CPI. Consistent with this, *CST3* gene expression was significantly elevated in the ‘immunologically- quiet’ C5 TCGA immune subtype (p<1e-10, Tukey’s test against all other subtypes, Figure S8a), which is characterized by the highest macrophage and lowest lymphocyte abundance. To investigate whether dynamically increased *CST3* gene expression would be associated with resistance to CPI, we reviewed paired pre- and post-treatment tumor biopsy scRNA-seq from patients (n=8) with metastatic basal cell carcinoma (BCC), treated with anti-PD-1^77^. Patients were split into responders (n=3) and non-responders (n=5, Figure S8b) according to radiological response. Pre-treatment *CST3* expression in macrophages, dendritic cells (DCs), cancer-associated fibroblasts (CAF) and tumor clusters did not predict CPI responsiveness (p>0.05, paired t-test). However, we observed evidence for significant dynamic *CST3* upregulation in CAFs and DCs in non-responder patients (p<0.05, paired t-test, Figure S8c-f).

We considered that the CyC-production PGS could reasonably capture the capacity to dynamically regulate secretion of CyC, and so predict failure of CPI. To estimate CyC-production PGS in patients treated with CPI, we collated 8 published cohorts of cancer patients treated with anti-PD-1, anti-PD-L1 or anti-CTLA-4 therapies with available germline exome sequencing (termed panIO cohort, Figure S6b, Table S2a). 685 patients with European ancestry passed quality control for inclusion (cohort characteristics summarized in Table S2b). Following imputation of common variants, the exome-wide CyC-production PGS was scored in each patient. Using multivariate Cox regression adjusted for sex, genetic ancestry and tumor type, we demonstrated that CyC-production PGS was associated with significantly worse progression-free survival (HR=1.29, p=0.0005) and worse overall survival (HR=1.09, p=0.10, Figure 6h). Using logistic regression with the same covariates, we further demonstrated that the PGS was associated with significantly reduced odds of durable clinical benefit (OR=0.78, p=0.003, Figure 6h). This latter effect was broadly consistent in each tumor type (Figure 6i). Altogether, these findings suggest that increased intratumoral CyC production may make a substantial contribution to failure of cancer immunotherapy, and that this effect may be mediated by recruitment of TREM2+ macrophages.

## Discussion

This work firmly establishes a direct link between GC signaling, CyC and Trem2+ macrophages. We demonstrate CyC’s biological and clinical relevance using a combination of genetic analyses, *in vitro*, *in vivo*, experimental medicine approaches, and clinically relevant prognostic and predictive studies. While the focus on human datasets allowed us to investigate clinically-relevant questions, we acknowledge that many of the analyses presented are limited by their associative nature. Although associations between measured CyC and clinical outcomes have the potential to be confounded in multiple directions, we agree with others that associations between patient-level polygenic scores and outcomes are substantially more robust, with the potential to capture causal associations^78^. In addition, we performed all PGS analyses in either a held-out validation cohort (for UKB) or an independent non-overlapping cohort (TCGA, panIO) to mitigate against the risk of overfitting. Thus, we argue that our findings make contributions to two gaps in our knowledge.

Firstly, estimation of renal function plays a central role in routine clinical practice by defining disease states, capturing acute systemic illness, and informing optimal medication dosing. Therefore, the relative strengths and weaknesses of CyC as a marker of renal function have substantial clinical relevance. It has been reported previously that CyC performs well as a renal function marker in healthy patients, but that its performance deteriorates in patients with acute disease and patients who receive GC agonists such as prednisolone, for example, renal transplant patients^54^. Our findings provide context for seemingly contradictory studies^15, 79^ measuring the effect of GC treatment on CyC levels by demonstrating that GC-inducible CyC secretion is context-dependent, and cannot be detected systemically in healthy mice and humans. One explanation for this context-dependency would be that an inflammatory stimulus would drive differentiation of monocytes to macrophages, in turn upregulating GR and enabling GC-dependent gene programs^61^, such as GC-inducible expression of *CST3*. Such a regulatory system would function to precisely tune the GC response program to minimize the off-target effects of GCs, which are well-recognized in clinical practice^80^. We recognize that other non-macrophage populations, such as dendritic cells, might secrete CyC in a GC-responsive manner. In the context of inflammatory diseases, such as COVID-19, we demonstrate that dexamethasone treatment induces detectable increases in systemic CyC levels and have recently reported that CyC levels are dynamically regulated in COVID-19 patients and associate with in-hospital mortality (2^nd^ manuscript in preparation). Altogether, this raises the question of whether CyC is a useful marker of renal function in patients with significant inflammatory disease, for whom estimation of kidney function is likely to be especially important.

Secondly, despite the widespread adoption of exogenous GCs as a treatment for inflammatory conditions and for treating autoimmune adverse effects of CPI, the exact upstream mechanisms by which GCs cause immunosuppression remains elusive^80^. Our findings, together with work from others, suggest that CyC qualifies as a candidate effector of GC-induced immunosuppression: CyC is biologically active as a potent cysteine protease inhibitor^12^, CyC secretion can be induced by GC agonists in inflammatory macrophages and this process is co-opted by cancer cells, for which the immune system is a dominant selection pressure^81^. Furthermore, CyC levels are highest in cerebrospinal and seminal fluid^12^, which may even suggest a role in immune privilege. We recognise that the GC effect is not exclusive to CyC. In support of this model, cancer cell-intrinsic CyC-knockout attenuates tumor growth kinetics and proliferation in immunocompetent mice, consistent with evidence that germline *CST3* knockout abrogates metastasis *in vivo*^82^. Using single-cell omics and population genetics, we propose a model in which CyC directs the recruitment of immunosuppressive Trem2+ macrophages^75, 76^, which in turn promote failure of cancer immunotherapy. However, future work has to determine the precise molecular mechanism by which this sequence occurs. One potential link is Apolipoprotein E (ApoE), which is known to be secreted by cancer cells^83^ and is a high-affinity ligand for the Trem2 receptor (Kd = 6 nM)^84^. Ligation of the Trem2 receptor by ApoE is sufficient to promote phagocytosis in TREM2-expressing microglial (brain-resident macrophage) cells, and in turn activates *Apoe* RNA expression^85^, suggesting the existence of an autocrine positive feedback loop. Given that ApoE can be proteolytically processed in a range of contexts^86^ and CyC is a potent protease inhibitor, it is conceivable that CyC might act to regulate ApoE availability in the tumor microenvironment, thereby regulating recruitment and proliferation of Trem2+ macrophages. Furthermore, evidence that M2-like macrophages appear to modulate the trans-eQTL association at rs2749527 between cortisol and *CST3* expression, but also express TREM2^75, 87^ suggests the existence of a single autocrine loop driving GC-induced expansion of TREM2+ macrophages. In support of connectivity between CyC, ApoE and TREM2, Trem2-knockout mice have accelerated amyloid burden in mouse models of Alzheimer’s disease (AD)^88^, TREM2 R47H mutations impair ApoE binding^84^, and increase the risk of human AD^89^, while CyC knockout is associated with reduced amyloid burden^90^. Consistent with a direct immunosuppressive function of CyC, we demonstrate that germline predisposition to CyC production is significantly associated with substantial remodeling of the intertumoral immune landscape and failure of cancer immunotherapy. The evidence that CyC-production PGS predicts failure of immunotherapy requires experimental confirmation that is beyond the scope of this present study. If confirmed, it would suggest that a combination of PD-1/PD-L1 blockade and CyC inhibition should be explored as a putative therapeutic approach in patients who do not respond to CPI.

## Data Availability

Due to the data use agreements for the datasets analyzed in this manuscript, we are unable to directly share or distribute any patient-level data except for COVID-19 patient scRNA-seq (reposited at https://doi.org/10.6084/m9.figshare.14330795.v13, filename 'covid.combined_final.CD14.Mono.Robj'). All summary statistics are published alongside the study, and polygenic scores will be reposited on PGS Catalog on the acceptance of the peer-reviewed manuscript. To facilitate dataset requests from applicable data use committees, we provide all accession codes for all datasets relating to this manuscript in the Methods and in Table S1. UK Biobank data can be requested through the application process detailed at https://www.ukbiobank.ac.uk/.
Single-cell RNA sequencing from mouse tumors will be deposited on SRA on acceptance of a peer-reviewed manuscript, the processed Seurat matrix is available from Figshare (https://doi.org/10.6084/m9.figshare.20063402). Primary data for in vitro and in vivo experiments will be published on acceptance of a peer-reviewed manuscript. All scanned slides will be made available through the CSHL data repository on acceptance of a peer-reviewed manuscript.
Code to reproduce core computational and statistical analyses has been reposited on Github at https://github.com/Janowitz-Lab/cystatinc.
Where data use agreements allow, additional information required to reanalyze the data reported in this paper is available from the lead contact upon request.

https://doi.org/10.6084/m9.figshare.20063402

## Acknowledgements

This work was conducted using the UK Biobank resource under application number 58510. All data access relating to this manuscript was approved by the institutional IRB at Cold Spring Harbor Laboratory, which determined that does not meet the definition of human subject research under the purview of the IRB according to federal regulations. We thank all patients and their families who have volunteered to participate in clinical research, without whom this study would not have been possible. T.J. acknowledges funding from Cold Spring Harbor Laboratory and grants from the Simons Foundation, Mark Foundation (20-028-EDV), and Nancy Gay Foundation (58391101). HVM receives funding from the Simons Center for Quantitative Biology at Cold Spring Harbor Laboratory. S.K. was supported by the Starr Centennial Scholarship at the Cold Spring Harbor Laboratory School of Biological Sciences. M.F. is supported by the “la Caixa” Foundation (ID 100010434) in the framework of the “La Caixa” Fellowship Program under agreement LCF/BQ/AA18/11680037. B.R.W. is supported by a Wellcome Investigator Award (107049/Z/15/Z). S.B. was supported by a Medical Research Council PhD studentship. Computational analyses were performed with assistance from the US National Institutes of Health Grant S10OD028632-01. L.G.N.A. received Alberta Innovates and Killam doctoral scholarships. S.S. received CIHR Vanier, Alberta Innovates and Killam doctoral scholarships. GJ acknowledges funding from Swedish Research Council (Project 2015-02561 and 2019-01112). Histological processing was performed by the CSHL Tissue Imaging Shared Facility with assistance from the Cancer Center Support Grant 5P30CA045508

## Author contributions

S.O.K., H.V.M. and T.J. conceived and designed the study. S.O.K. performed statistical and computational analyses, with support from T.H. S.B. performed eQTL analyses in the STARNET cohort. H.L. contributed to data curation of clinical cohorts. B.D. performed *in vitro* experiments, while S.O.K., N.M., B.D. and M.F. performed *in vivo* experiments. C.R. performed single-cell library preparation. S.K. performed single-cell analysis, with support from Y.J.R.A.R and guidance from J.P. D.C. performed analyses of human clinical trial specimens under supervision of G.J. S.S., N.R., B.Y., J.B., A.D., L.G.N.A., P.T., M.R., F.K. and V.D. performed proteomic analyses of COVID-19 patient specimens. Q.G. optimized and executed histological staining. Scanned slides were computationally analyzed by D.L. under supervision of V.H.K. B.W. provided input for interpretation of results and direction for further analyses. H.V.M and T.J. supervised the project and guided all data analyses. S.O.K., H.V.M. and T.J. wrote the manuscript with input from all authors. All co-authors approved the final version of the manuscript.

## Figures and figure legends

**Figure S1.**
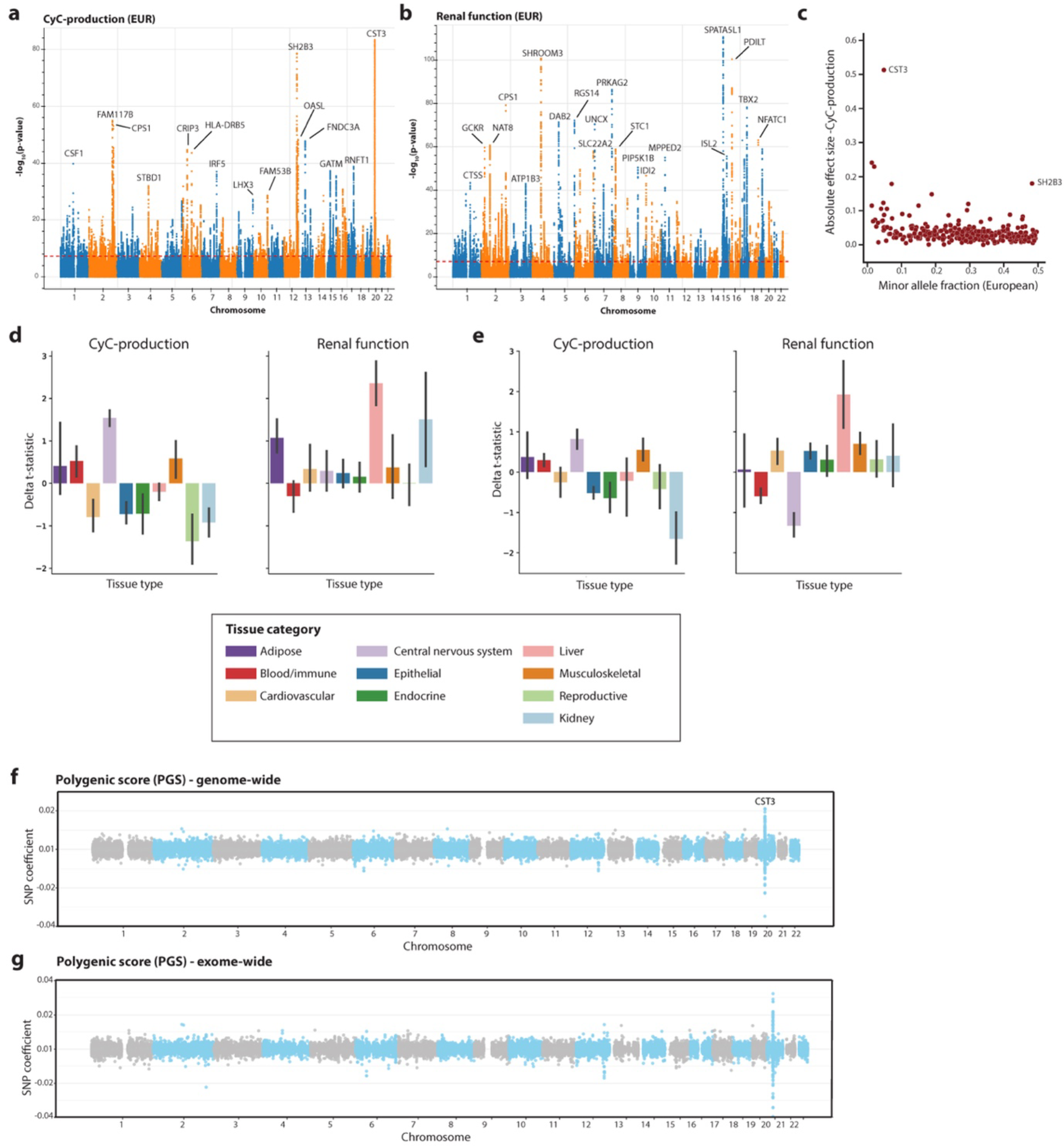
Summary statistics from (a) Cystatin C (CyC)-production and (b) Renal function latent traits in European UKB subjects, displayed a Manhattan plot. Loci with a p-value less than 1e-30 are annotated with gene name from OpenTargets V2G pipeline. (c) Relationship between effect size and minor allele frequency in CyC-production trait, annotated with outlier loci. Partitioned heritability analysis across multiple tissue types derived from (d) gene expression and (e) chromatin accessibility data. Delta t-statistic refers to change in enrichment t-statistic between measured eGFR-CyC summary statistics and latent CyC-production or renal function statistics. Errors bars signify 95% confidence interval. Coefficients for each SNP included in polygenic scores (PGS) generated using (f) HapMap SNPs (n=1,000,000) or (g) HapMap SNPs that can be reliably imputed from exome (n=300,000) sequencing data. CST3 locus on chromosome 20 is annotated.

**Figure S2.**
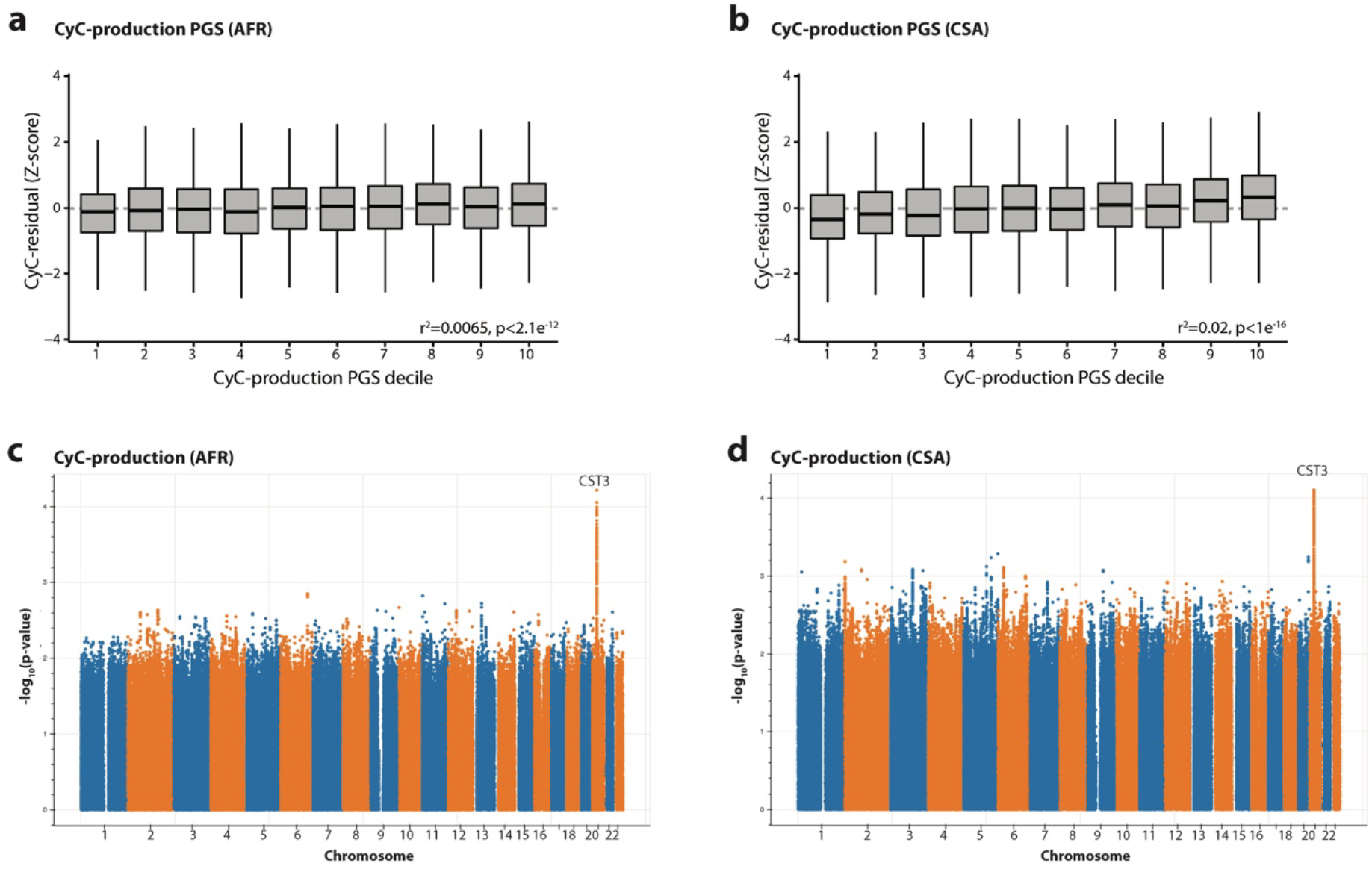
Trans-ancestral portability of CyC-production polygenic score (PGS) derived in UK Biobank European training set (n=381,764) applied to subjects of (a) African (AFR, n=8152) and (b) Central and South Asian (CSA, n=9845) genetic ancestry. Boxplots show median (central line) with interquartile range (IQR, box) and extrema (whiskers at 1.5× IQR). Summary statistics from CyC-production latent trait in (c) AFR and (d) CSA genetic ancestry cohorts, derived from GWAS for eGFR-CyC and eGFR-creatinine followed by structural equation modeling. Results displayed as Manhattan plot; no loci reached genome-wide significance in latent trait analysis.

**Figure S3.**
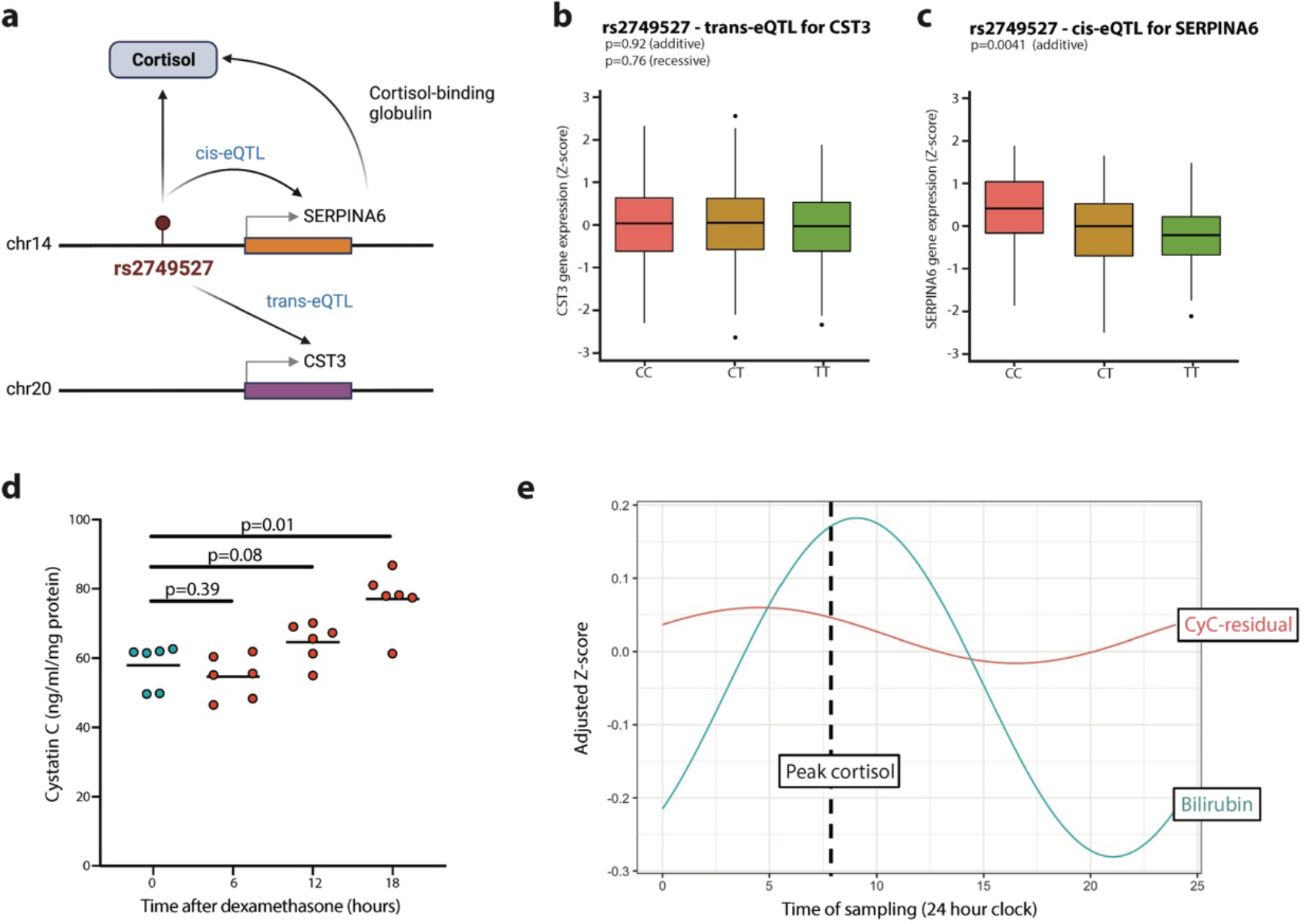
(a) Role of single genetic instrument, rs2749527, as a trans-eQTL for *CST3* on chromosome 20 (in visceral adipose fat) and as a cis-eQTL for *SERPINA6* on chromosome 14 (in liver), which codes for cortisol-binding globulin, that is also significantly associated with morning plasma cortisol. Association analysis between rs2749527 and both (b) *CST3* gene expression in visceral adipose fat (VAF, n=381) and (c) *SERPINA6* gene expression in liver (n=175) in GTEX cohort (EUR ancestry). Boxplots show median (central line) with interquartile range (IQR, box) and extrema (whiskers at 1.5× IQR). Outliers beyond 1.5× IQR are shown as dots. (d) Extracellular cystatin C concentrations in A549 cells normalized to cellular protein content after 0-, 6-, 12- or 18-hour treatment with 100nM dexamethasone. Each timepoint comprises 6 biological replicates. (e) Diurnal variation in CyC-residual and bilirubin (known to correlate with plasma cortisol) derived from cosinor regression in UKB cohort. Adjusted Z-score refers to Z-scoring stratified by age and gender.

**Figure S4.**
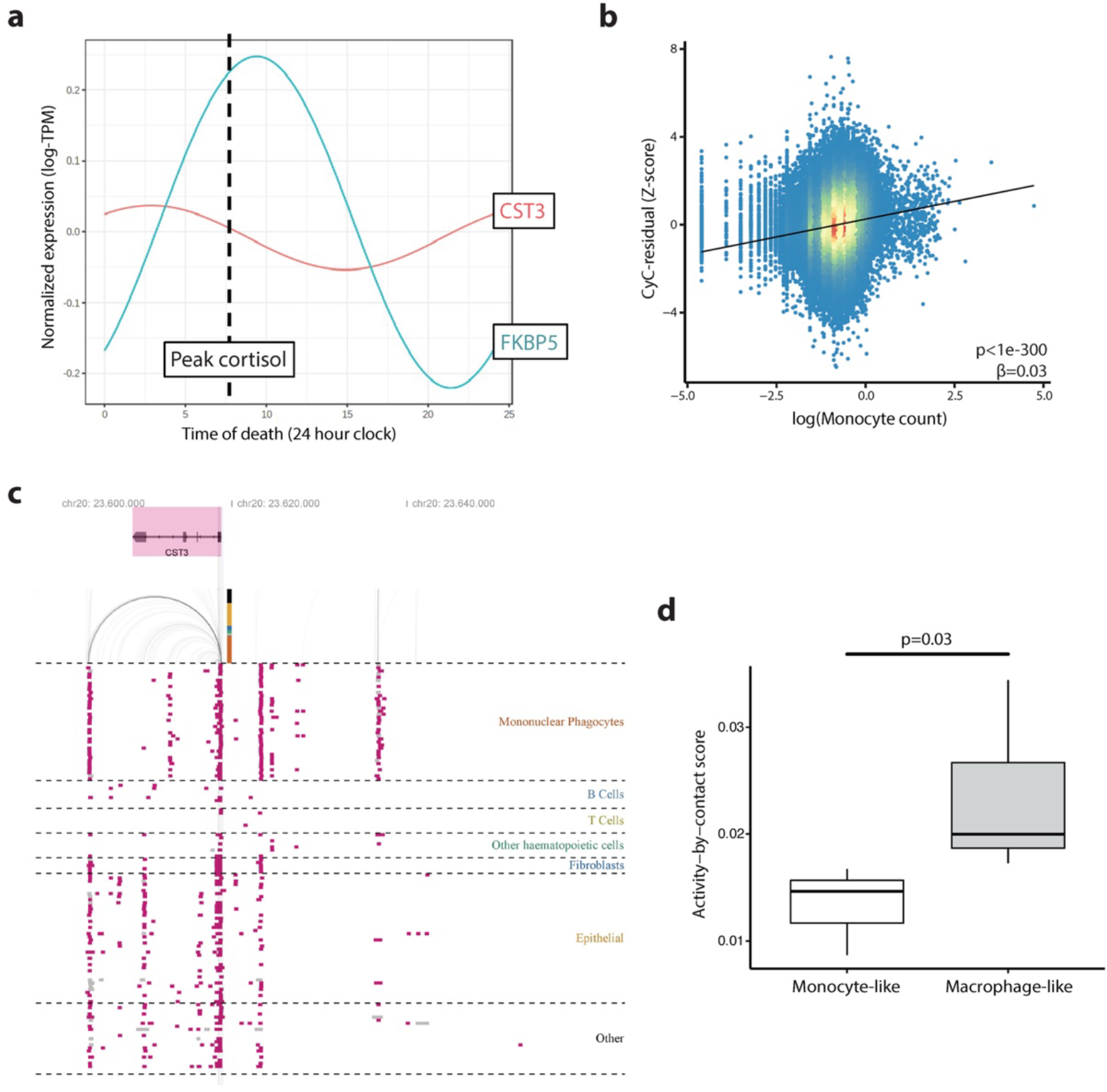
(a) Diurnal variation in *CST3* and *FKBP5* (canonical glucocorticoid response gene) derived from cosinor regression in GTEX spleen cohort, using time of death for each GTEX donor. (b) Significant positive correlation between logarithm of monocyte count and Z-score cystatin C (CyC)-residual in UKB cohort. Significance refers to multivariate regression including age, sex and body mass index (BMI). (c) Visualization of predicted enhancer elements at *CST3* locus from activity-by-contact (ABC) model^93^, showing distal enhancer element acting on cystatin C. Each row corresponds to a cell line-treatment pair and epithelial grouping includes cancer cell lines. (d) ABC model scores for distal enhancer in THP-1 cells - with (macrophage-like) or without (monocyte-like) PMA treatment.

**Figure S5.**
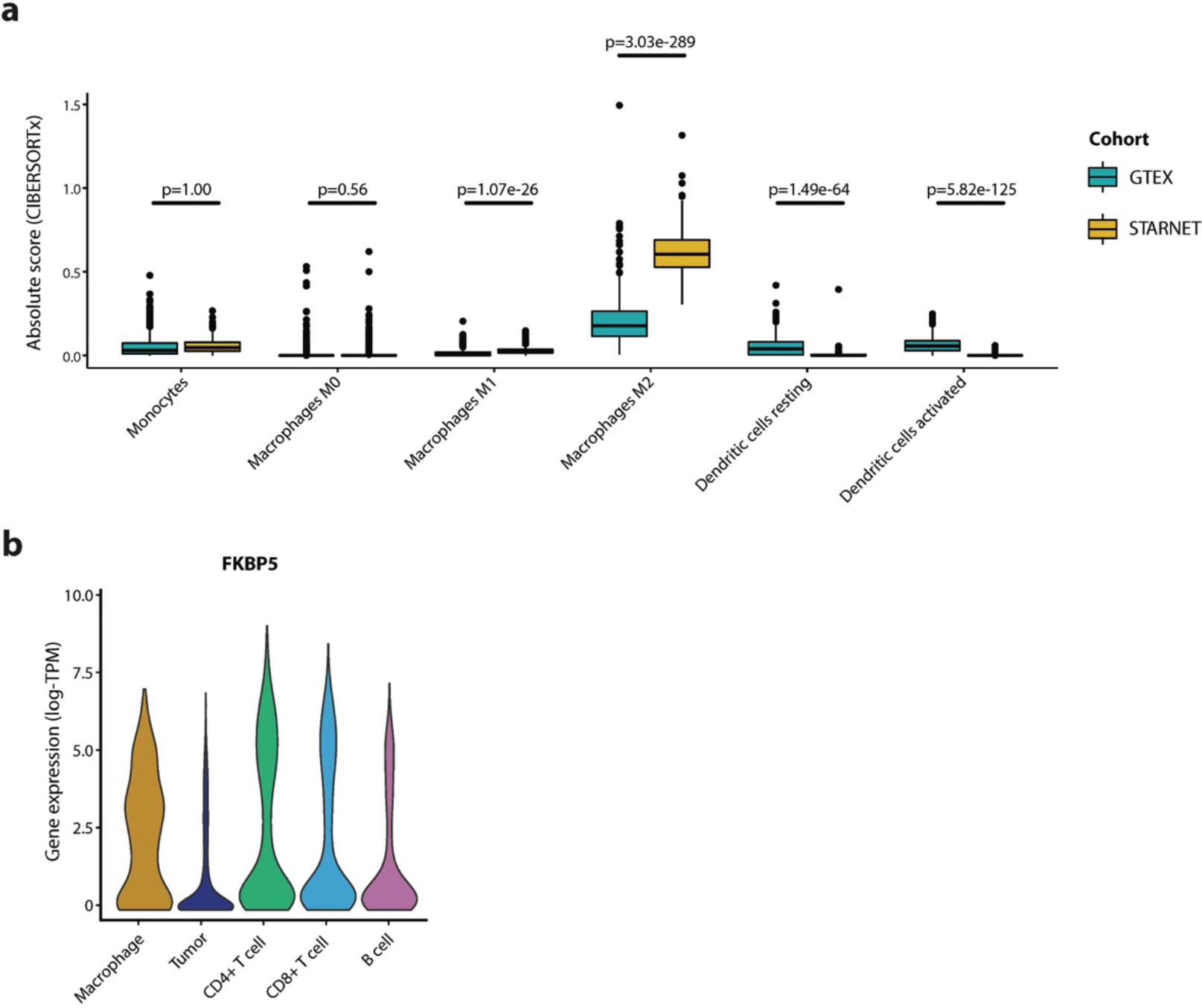
(a) Inferred absolute myeloid cell composition from CIBERSORTx analysis (absolute mode) applied to visceral adipose tissue (VAF) from STARNET and GTEX cohorts. p values refer to t-tests with Bonferroni correction. While units are comparable between cell types and samples, they do not refer to an absolute cell fraction. Boxplots show median (central line) with interquartile range (IQR, box) and extrema (whiskers at 1.5× IQR). Outliers beyond 1.5× IQR are shown as dots. Marker genes used to define M0-like, M1-like (such as *CCL19*) and M2-like (such as *CCL18*) macrophages are described in the CIBERSORT manuscript^94^. (b) Single-cell *FKBP5* (canonical glucocorticoid receptor target) gene expression in each cell cluster in melanoma tumors (n=12) from Jerby-Anon et al^95^. Clusters defined by correlation to reference PBMC data^91^, with unclassified cells that exhibit detectable clonal copy number variation classified as tumor.

**Figure S6.**
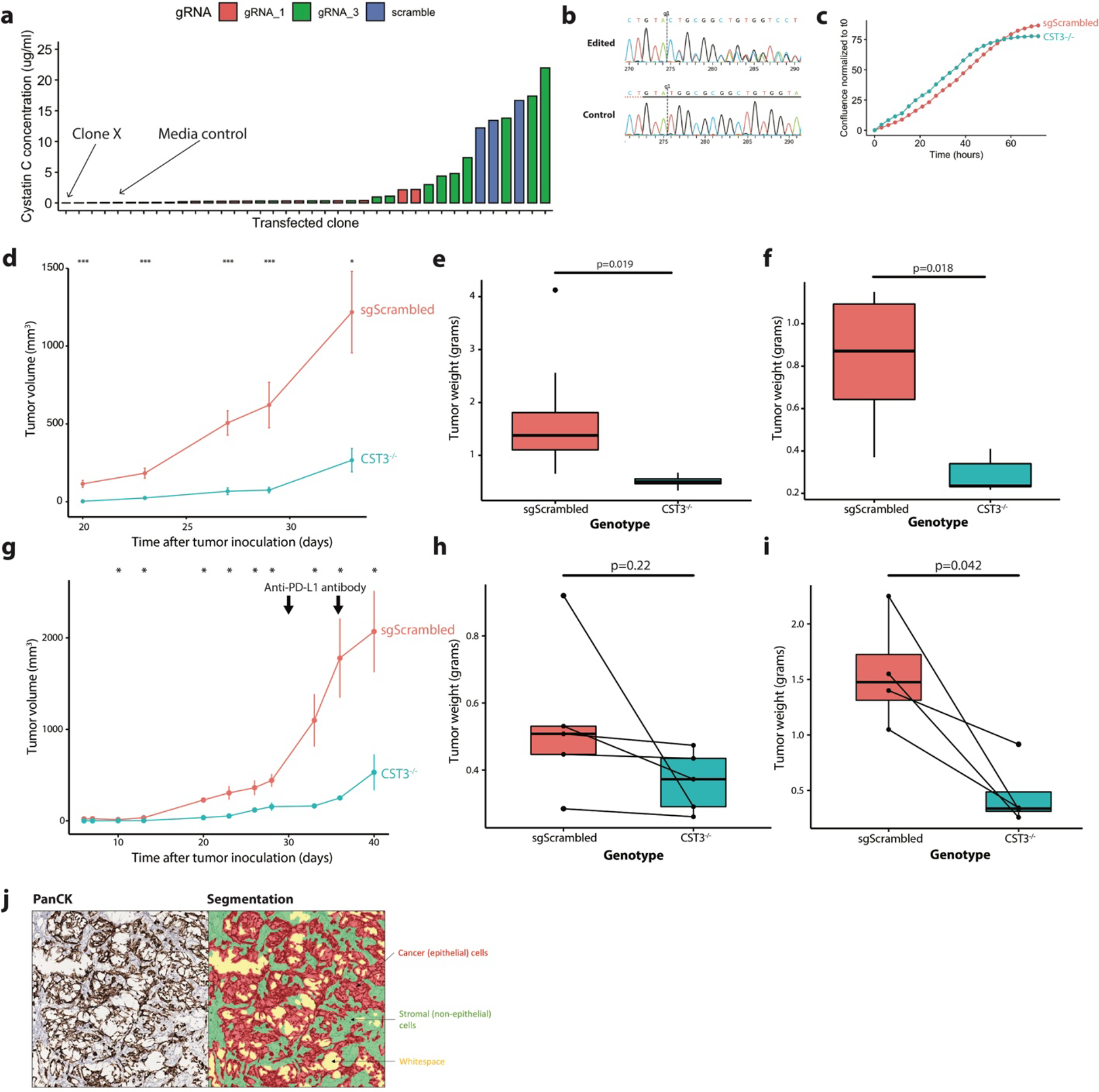
(a) Extracellular cystatin C (CyC) concentrations measured by ELISA in monoclonal cell populations derived from transfection of Mm1 cells with guide RNAs (gRNAs) specific to the *CST3* gene locus (gRNA_1 and gRNA_2, plus sgScrambled control). Clone X was selected on the basis of lowest extracellular CyC concentration. (b) Sanger sequencing trace showing high editing efficiency (>97%) at the predicted binding site for gRNA_1 in clone X. (c) Cell confluence kinetics for sgScrambled and *CST3*^-/-^ (clone X) cells normalized to confluence at the start of the experiment. (d) Tumor growth curves (mean and standard error of the mean) for an independent replication with single-flank sgScrambled (n=10) and *CST3*^-/-^ (n=10) tumors, 200,000 cells inoculated in right flank (cohort B). Endpoint tumor weights at endpoint for (e) cohort A and (f) cohort B. (g) Tumor growth curves (mean and standard error of the mean) for an independent replication bi-flank paired sgScrambled (n=4) and *CST3*^-/-^ (n=4) tumors, 100,000 cells inoculated in both flanks (cohort D). Mice received two doses of anti-PD-L1 antibody. Endpoint tumor weights at endpoint for (h) cohort C and (i) cohort D. (j) Representative Ki67 immunohistochemistry annotated with automated segmentation of epithelial and non-epithelial compartments.

**Figure S7.**
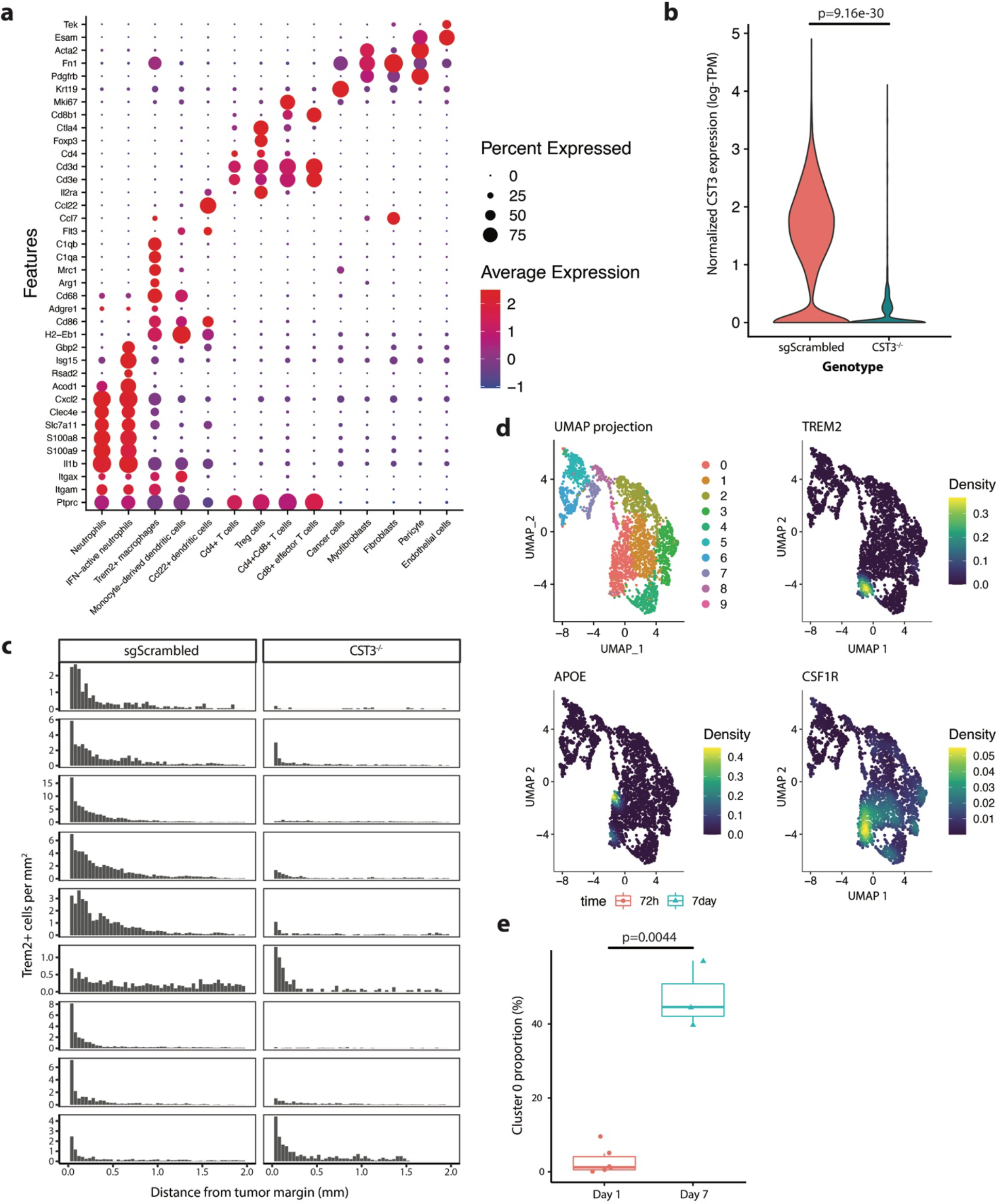
(a) Dotplot of marker gene expression (marker selection detailed in Table S7) used to define cell cluster identity in scRNA-seq of Mm1 tumors. Dot color signifies mean expression (log- TPM) while dot size signifies the proportion of each cell population that have detectable expression of each gene. (b) Violin plot showing cancer cell compartment-specific *CST3* gene expression in sgScrambled and CST3^-/-^ tumor samples. P-value refers to a pseudobulk comparison. (c) Histogram summarizing number of Trem2+ cells per mm^2^ in each 40μm window from the tumor margin in paired sgScrambled and CST3^-/-^ tumor sections, each row corresponds to a mouse (n=9) with tumors inoculated in each flank. (d) UMAP plots of re-clustered CD14+ monocytes (n=10 clusters) from ICU-admitted COVID-19 patients^64^ annotated with the *Nebulosa*^73^ kernel function corresponding to TREM2, APOE and CSF1R gene expression; indicating that cluster 0 is enriched with Trem2+ monocytes. (e) Proportion of cluster 0 monocytes at day 1 (within 72 hours of admission) and day 7 in ICU-admitted patients with COVID-19 who were treated with dexamethasone. p-value is adjusted p-value from linear model of logit-transformed proportions.

**Figure S8.**
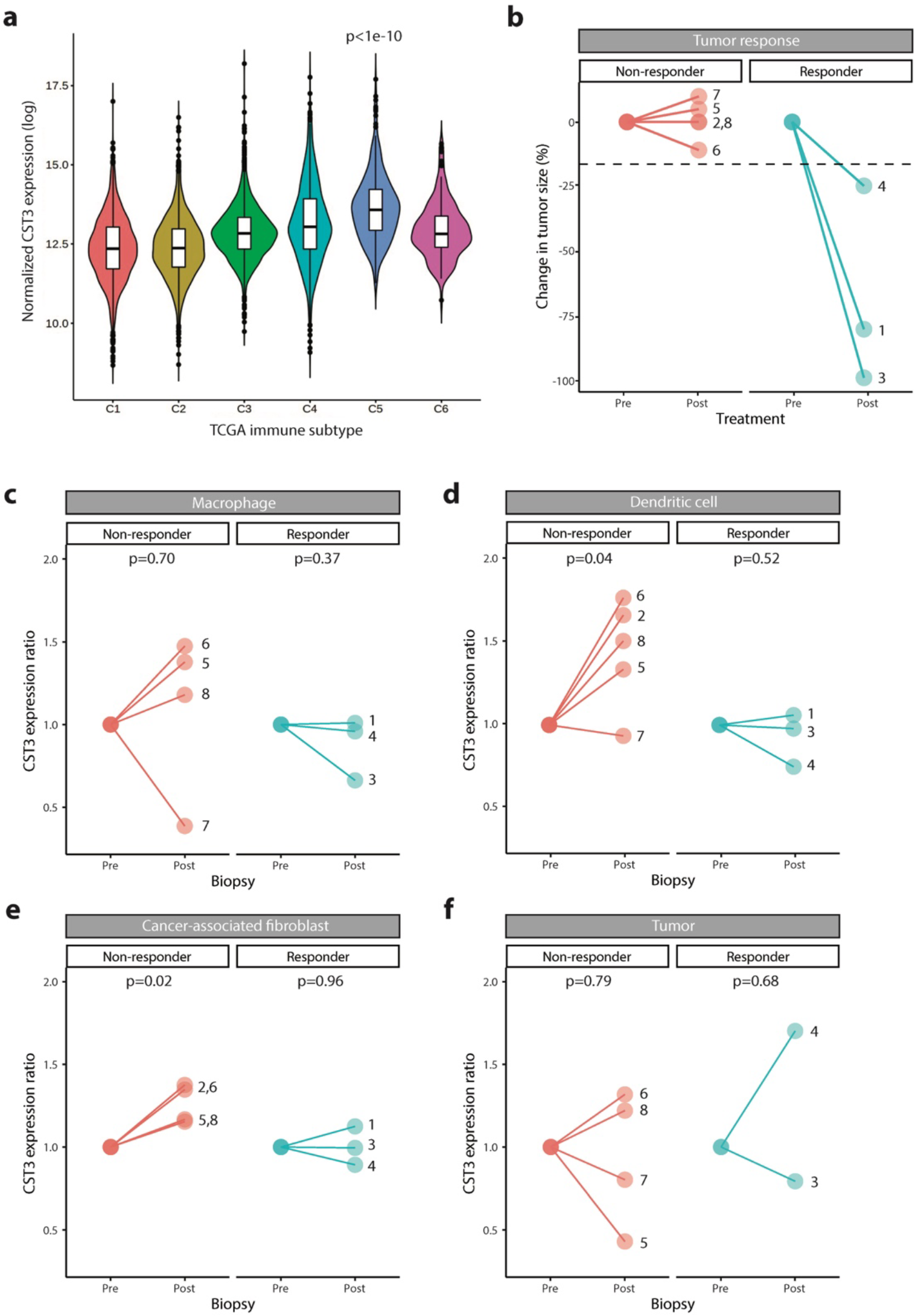
(a) *CST3* gene expression (log-normalized) in each TCGA immune subtype (Thorssen et al^96^). Subtype 5 corresponds to ’immunological quiet’ subtype, characterized by reduced lymphocyte and increased M2 macrophage responses. Significance level refers to one-way ANOVA with post-hoc Tukey’s test. (b) Summary of radiological response data for patients with basal cell carcinoma (BCC, n=8) derived from Yost et al^97^. Response is nominally defined as tumor regression >25%. Numbers refer to patient IDs. Ratio of cluster-specific pseudo-bulk *CST3* gene expression (log-TPM) in paired biopsy samples pre/post anti-PD-1 treatment, segregated according to clinical response, for (c) macrophage, (d) dendritic cell, (e) cancer-associated fibroblast (CAF) and (f) tumor subsets. Uncorrected P values refer paired t-tests.

## STAR Methods

### Key resources table

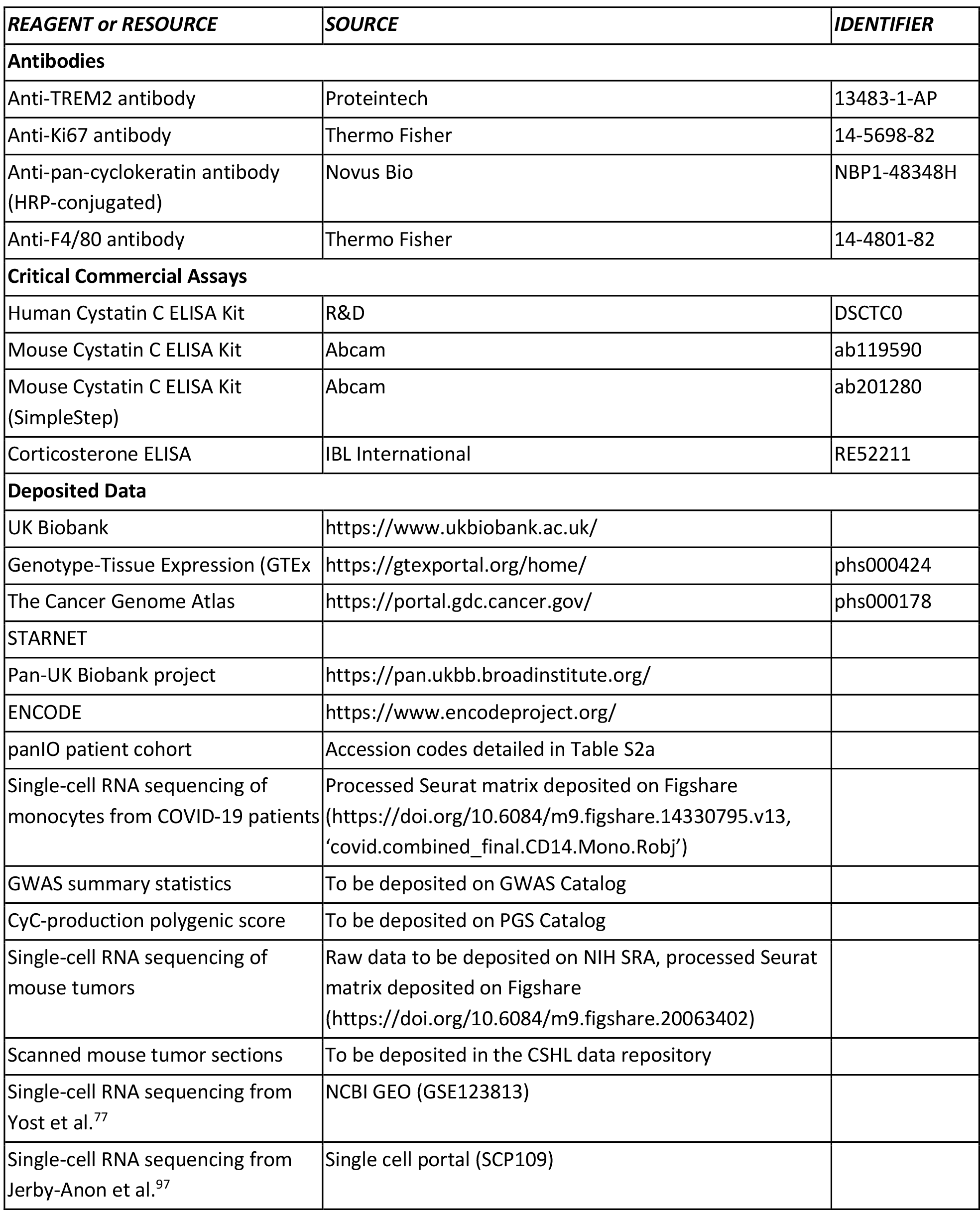
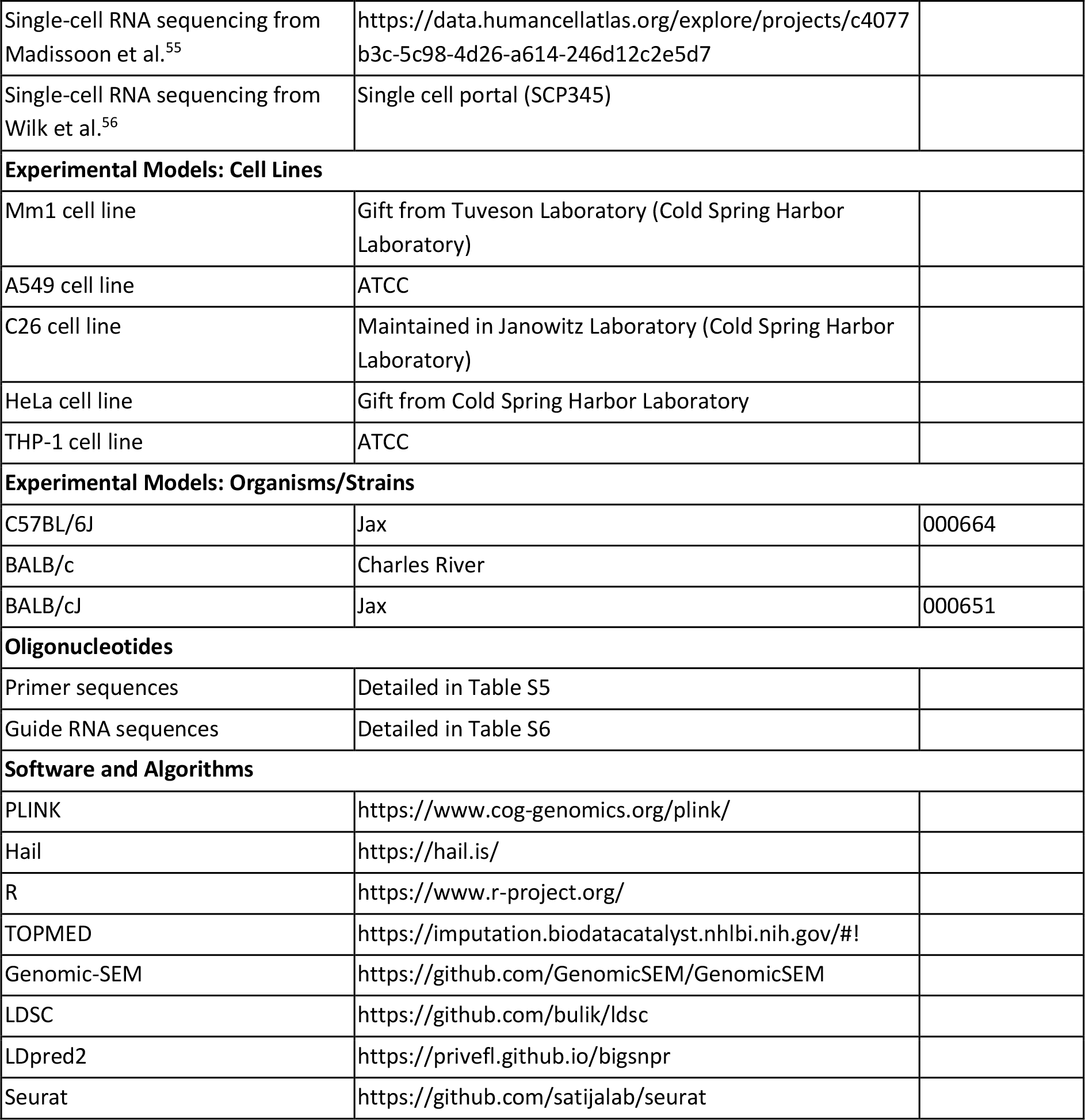

### Resource Availability

#### Lead Contact

Further information and requests for resources should be directed to the Lead Contact, Tobias Janowitz.

#### Materials Availability

CyC^-/-^ Mm1 cell line that was generated as part of this study is available from the lead contact with a completed material transfers agreement.

#### Data and Code Availability

- Due to the data use agreements for the datasets analyzed in this manuscript, we are unable to directly share or distribute any patient-level data except for COVID-19 patient scRNA-seq (reposited at https://doi.org/10.6084/m9.figshare.14330795.v13, filename ‘covid.combined_final.CD14.Mono.Robj’). All summary statistics are published alongside the study, and polygenic scores will be reposited on PGS Catalog on the acceptance of the peer-reviewed manuscript. To facilitate dataset requests from applicable data use committees, we provide all accession codes for all datasets relating to this manuscript in the Key Resources Table and in Table S2a. UK Biobank data can be requested through the application process detailed at https://www.ukbiobank.ac.uk/.
- Single-cell RNA sequencing from mouse tumors will be deposited on SRA on acceptance of a peer-reviewed manuscript, the processed Seurat matrix is available from Figshare (https://doi.org/10.6084/m9.figshare.20063402). Primary data for *in vitro* and *in vivo* experiments will be published on acceptance of a peer-reviewed manuscript. All scanned slides will be made available through the CSHL data repository on acceptance of a peer-reviewed manuscript.
- Code to reproduce core computational and statistical analyses has been reposited on Github at https://github.com/Janowitz-Lab/cystatinc.
- Where data use agreements allow, additional information required to reanalyze the data reported in this paper is available from the lead contact upon request.

### Experimental model and subject details

#### Cell line models

Human lung carcinoma cell line A549 was purchased from ATCC (CCL-185). Human cervical cancer cell line HeLa was obtained from Cold Spring Harbor Laboratory. Human acute monocytic leukemic cell line THP-1 was purchased from ATCC (TIB-202). Mm1 cells were a gift from D. Tuveson (Cold Spring Harbor Laboratory, NY), and are derived from a liver metastasis in the KPC model of pancreatic ductal adenocarcinoma^69^. A549, HeLa and Mm1 cell lines were cultured in DMEM supplemented with 10% FBS and 1% penicillin-streptomycin. THP-1 and C26 cells were cultured in RPMI supplemented with 10% FBS and 1% penicillin-streptomycin. Macrophage-like differentiation in THP-1 cells was induced by treatment with 50nM PMA (Sigma) for 48 hours, before replacement with PMA-free media and recovery for 24 hours prior to treatment. Cell viability was checked by trypan blue method and was consistently above 95% prior to seeding. All cell lines were cultured at 37°C in 5% CO2. Dexamethasone and PMA (phorbol 12-myristate 13-acetate) were purchased from Sigma-Aldrich. DMEM and RPMI cell culture media, fetal bovine serum (FBS), penicillin/streptomycin (P/S) and Dulbecco’s phosphate-buffered saline (DPBS) were purchased from Gibco

#### Mouse models

Wild-type BALB/c mice obtained from Charles River Laboratories (for C26 model of cancer progression) and Jax (for dexamethasone treatment); and wild-type C57BL/6J obtained from Jax. All mice examined as part of this study were male as C2 and Mm1 lines were isolated from male mice. Mice were allowed to acclimatize for 7 days from arrival in the Cold Spring Harbor Laboratory animal facility. All animal experiments and care were performed in accordance with the Cold Spring Harbor Laboratory (CSHL) Institutional Animal Care and Use Committee (IACUC) and the National Institutes of Health Guide for the Care and Use of Laboratory Animals. Mice were kept in specific pathogen-free conditions on a 24 hour 12:12 light-dark cycle. Tumor samples were obtained by dissection of mice after euthanasia by cervical dislocation and tumor weights were routinely recorded. Plasma samples were obtained from tail bleeds and terminal cardiac bleeds. Tail bleeds were performed using a scalpel via tail venesection without restraint, and terminal bleeds were obtained at endpoint (cachexia) through exsanguination via cardiac puncture under isoflurane anesthesia. Samples were kept on ice at all times. Plasma samples were collected into heparin-coated capillary tubes to avoid coagulation and were processed as follows: centrifuge spin at 14,000 rpm for 5 minutes at 4°C, snap frozen in liquid nitrogen, and stored at -80°C.

#### Human studies

This study incorporates human subjects from three independent studies. All human subjects gave informed consent, and all studies were approved by the respective institutional review boards.

1. *Cohort 1.* This cohort has been reported previously^59^ and refers to a prospective, single-center, single-blind randomized crossover clinical trial that recruited 10 subjects (men and women) with primary adrenal insufficiency (Addison’s disease). The study was approved by the Ethics Review Board of the University of Gothenburg, Sweden (permit no. 374-13, 8 August 2013) and conducted in accordance with the Declaration of Helsinki. Written informed consent was obtained from all subjects before participation. The study was registered at ClinicalTrials.gov with identifier NCT02152553.
2. *Cohort 2.* This cohort has been reported previously^64^ and refers to a prospective study that recruited 14 patients with COVID-19 necessitating admission to ICU, of which 6 received dexamethasone treatment as part of their clinical course. All patients or their surrogate decision-makers gave informed consent for participation. This study was approved by the Conjoint Health Research Ethics Board at the University of Calgary (Ethics ID: REB20-0481) and is consistent with the Declaration of Helsinki.
3. *Cohort 3.* This cohort (Pa-COVID-19 study) has been reported previously^65^ and refers to a prospective observational cohort study at Charité Universitätsmedizin Berlin. Patients with a PCR-confirmed diagnosis of SARS-CoV-2 infection were eligible for inclusion in the study. The Pa-COVID-19 study is carried out according to the Declaration of Helsinki and the principles of Good Clinical Practice (ICH 1996) where applicable and was approved by the ethics committee of Charité- Universitätsmedizin Berlin (EA2/066/20).

### Method details

#### *In vitro* glucocorticoid treatment

For all experiments, cells were plated in 6 well plates, at a density of approximately 500,000 cells/ml. Cells reached confluence on day one or day two after being seeded. For time course experiments, cells were seeded and harvested at the same time, with the only variable being the duration of treatment with 100nM dexamethasone (varied between 0 and 18 hours), with 0-hour treatment acting as the control. For single-timepoint experiments, cells were treated with either 100nM dexamethasone (Sigma) or 0.01% ethanol for 18 hours prior to harvesting. For each experiment all samples were harvested concurrently.

For quantification of extracellular CyC, cell supernatant was collected at harvesting, spun at 10000 x g for 5 minutes to remove debris, and analyzed by ELISA (Human Cystatin C ELISA Kit, R&D Systems; Mouse Cystatin C ELISA Kit, Abcam/ab119590), with each sample profiled in duplicate. For quantification of cellular protein content, cells were washed with DPBS and ice-cold RIPA buffer with protease and phosphatase inhibitors (Thermo Fisher) was added to each well. The cell lysate was passed through a 25G syringe for homogenization and spun for 10000 x g for 15 minutes, at 4°C. The supernatant from the spun-down lysate was then stored at -80 for later analysis. BCA assay was performed on the cell lysate, with each sample profiled in duplicate. Normalized extracellular CyC concentrations were determined by dividing the ELISA-derived CyC concentration by the BCA-derived cellular protein content.

For quantitative real-time polymerase chain reaction (RT-PCR), RNA was extracted using the RNeasy Mini Kit (Qiagen) and reverse transcribed using SuperScript IV VILO Master Mix (Thermo Fisher) according to the manufacturer’s protocol. Four housekeeping genes (GUSB, PPIA, RPL15, RPL19) with minimal variation on GC treatment were selected on the basis of a literature review^98^ and differential expression analysis in ENCODE RNA-seq data (accession ENCSR897XFT), implemented in edgeR. Primers were designed using NCBI Primer-BLAST, with exon-spanning primers designed where possible (primer sequences detailed in Table S5). PCR was performed using the PowerTrack SYBR Green Master Mix (Thermo Fisher) using the QuantStudio 6 Flex (Thermo Fisher) instrument, using a 10μl reaction volume in technical triplicate according to the manufacturer’s protocol. The threshold cycle was determined by the Second Derivative Maximum method and the expression of each target was normalized relative to the geometric mean of four endogenous controls.

#### *In vivo* glucocorticoid treatment

Wild-type BALB/c and C57BL/6J were treated with a single dose of 20mg/kg dexamethasone given intraperitoneally (IP) at 9am. Dexamethasone 21-phosphate disodium salt (Sigma) was dissolved in PBS and filter sterilized prior to injection. Tail vein samples were taken 24- and 48- hours following IP dosing, and plasma levels of CyC were determined with Mouse Cystatin C ELISA Kit (ab119590), Abcam.

#### Glucocorticoid treatment in human subjects

Glucocorticoid treatment in human cohort 1 has been reported previously^59^. Briefly, subjects were randomized to a 22-hour treatment (commencing at 9am) with placebo (intravenous 0.9% saline) or near-physiological glucocorticoid treatment with intravenous hydrocortisone. During the GC exposure, hydrocortisone was administered at a varying dose of 0.024 mg/kg/hr between 9 AM and 12 PM (first day), 0.012 mg/kg/hr between 12 PM and 8 PM (first day), 0.008 mg/kg/hr between 8 PM and 12 AM (first day), and 0.030 mg/kg/hr between 12 AM and 7 AM (second day). After 2 weeks, subjects were given whichever treatment they had no yet received, as part of a crossover study design. Blood samples were collected in the morning of the second intervention day (6 AM) and plasma was isolated. Plasma CyC and creatinine were measured used validated clinical assays (creatinine: Alinity c Creatinine (Enzymatic) Reagent Kit; CyC: Gentian Cystatin C Immunoassay) at the laboratory of Sahlgrenska University Hospital in Gothenburg, Sweden.

#### CyC quantification in patients with COVID-19

For human cohort 2, serum samples were collected as specified timepoints (timepoint 1: within 72 hours of admission/referred to as day 1, and timepoint 2: 7 days after timepoint 1)^64^. ELISA-based serum cystatin C measurement was performed by Eve Technologies (Custom Human Kidney Injury Panel – Cystatin C). For human cohort 3, plasma sampling for plasma proteomics by mass spectrometry was performed three times per week subsequent to inclusion. Sample processing, mass spectrometry and data analysis were performed as described previously^22^, allowing for quantification of plasma CyC levels in 309 patients. Out of these patients, 131 had available paired serum creatinine for at least one timepoint, as well as clinical outcome data (COVID-specific mortality). For patients with at least one creatinine measurement, missing data were imputed with the most recent value. Plasma CyC levels were scaled by a factor of 300, so that the cohort mean was comparable to the mean serum CyC recorded in the UKB cohort (field 30720, units mg/L). For each patient, a creatinine-CyC (C2) ratio was calculated at each timepoint, using CKD-EPI eGFR equations with the race term set to 0.

#### *In vivo* model of cancer progression

Experiments with the C26 model were performed using 8-weeks old wild-type BALB/c male mice. Mice were inoculated subcutaneously in their right flank with the syngeneic C26 colorectal cancer cell line (2×10^6^ viable cells in 100μl RPMI vehicle) that induces cachexia. Prior to inoculation, C26 cells were dissociated with trypsin, followed by resuspension in FBS-free RPMI and counting of the viable cell concentration (trypan blue). C26-tumor bearing mice were termed pre-cachectic from 18 days post-inoculation and were defined as cachectic when their weight loss exceeded 15% from peak body weight. Plasma levels of CyC were determined with Mouse Cystatin C ELISA Kit (ab119590), Abcam. Corticosterone levels were quantified using Corticosterone ELISA (RE52211) from IBL International (TECAN).

#### Establishment of isogenic CyC^-/-^ cell line

Mm1 were transiently transfected with CRISPR plasmids (PX459, GenScript) encoding either a guide RNA (gRNA) specific to a coding region in mouse *Cst3* or a non-targeting (scrambled) gRNA. We tested two *Cst3*-specific gRNAs and one scrambled gRNA from a pre-validated database^99^. Guide RNA sequences are summarized in Table S6. Mm1 cells were seeded into 24-well plates with 50,000 cells per well and after 24 hours, they were transfected with 500ng plasmid using Lipofectamine 3000 (Thermo Fisher) according to the manufacturer’s protocol. We included a GFP-expressing plasmid to assess transfection efficiency. After 48 hours, the media was changed and replaced with DMEM media supplemented with 5μg/ml puromycin. After 72 hours, the media was replaced with DMEM media for 24 hours, followed by isolation of monoclonal populations by serial dilutions in a 96-well plate. To identify clones with CyC knockout, we measured CyC in the cell supernatant for each clone using the Mouse Cystatin C ELISA Kit (ab201280), Abcam. To verify the presence of truncating mutations in the *Cst3* coding region, we extracted genomic DNA from each clone (Qiagen DNeasy Blood and Tissue Kit) and performed targeted polymerase chain reaction (PCR) amplification and Sanger sequencing of the predicted gRNA binding sites. The editing efficiency was assessed using the Synthego ICE Analysis tool (https://ice.synthego.com/).

#### Characterization of isogenic CyC^-/-^ cell line

To compare the *in vitro* growth kinetics of isogenic sgScrambled and CyC^-/-^ cell lines, cells were seeded into 6-well plates with 200,000 cell per well, with three biological replicates per clonal cell line. Each well was scanned every 2 hours using an IncuCyte S3 Live Cell Analysis Instrument using the phase channel according to the manufacturer’s protocol. Cell confluence was estimated using the IncuCycte Cell-By-Cell analysis module, and was normalized to the first timepoint. The doubling time was estimated by fitting a model of log(time) as a function of confluence. To compare the *in vivo* growth kinetics of isogenic sgScrambled and CyC^-/-^ cell lines, mice were inoculated subcutaneously with 50,000-200,000 cells in the flank. For uni-flank experiments, mice were inoculated in the right flank; for bi-flank experiments, mice were inoculated in both left and right flanks, with the sgScrambled tumors on the left flank and the CyC^-/-^ tumor on the right flank. For tumor inoculation, Mm1 cells were dissociated with trypsin followed by resuspension in FBS-containing DMEM, counting of the viable cell concentration (trypan blue) and resuspension in sterile PBS. 10-20μl of PBS-suspended cell mixture was combined with an equal volume of Cultrex Reduced Growth Factor Basement Membrane Extract (3433-010-01, R&D Systems) on ice. Immediately prior to inoculation, the suspended cell mixture is thawed to room temperature and loaded into insulin syringes (328440, BD). Mice were monitored regularly until palpable tumors formed, after which point the longest and shortest dimensions of each tumor was measured every 3-4 days using calipers. For anti-PD-L1 treatment, mice were treated with 200μg of anti-PD-L1 monoclonal antibody (BioXCell, BP010) every 3 days, given intraperitoneally (IP). Unless otherwise stated, mice were sacrificed by cervical dislocation once tumors exceeded 20mm on one axis.

#### Single-cell RNA sequencing of mouse tumors

Tumors were finely minced at 4°C and transferred into tumor digestion medium containing collagenase/hyaluronidase and DNase I in RPMI 1640 with glutamine, then incubated on a shaker for 45 min at 37C and 300rpm. Freed cells were collected by passing through the dissociated tumor and media into a 70um cell strainer and quenching with FACS buffer at 4°C. Cells were spun down at 300g for 5 minutes at 4°C, the pellet resuspended in ice-cold ammonium chloride solution for 5 minutes and quenched with FACS buffer. Cells were spun down again and resuspended in FACS buffer. Viable cells were quantified by trypan blue method and samples were then subject to dead cell removal (EasySep Dead Cell Removal Kit, STEMCELL). Prior to library preparation, viability and cell number were re-assessed with a Countess II FL using AOPI (PN- CS2-0106-5mL, Nexcelom Bioscience). Single-cell RNA-seq libraries targeting 8,000 cells per sample were generated using the Chromium Next GEM Single Cell 3ʹ Reagent Kits v3.1 (PN- 1000121, 10x Genomics) according to the manufacturer’s instructions. Final libraries were sequenced to at least 25,000 reads per cell with the Illumina NextSeq 2000 and aligned with Cell Ranger (version 6.0.0, 10x Genomics) to the mm10 reference genome (refdata-gex-mm10-2020- A, 10x Genomics).

#### Tumor immunohistochemistry

Tumors were harvested and embedded in tissue molds containing OCT (Sakura) and frozen on dry ice prior to storage at -80°C. IHC staining were performed at CSHL Tissue Imaging Shared Facility. OCT embedded fresh tissue blocks were sectioned with Thermo #NX50 cryostat. 10μm thick sections were collected and mounted on positive charged glass slides (VWR superfrost plus micro slide) IHC slides were stained on DISCOVERY ULTRA IHC/ISH research platform (Roche) following standard protocols. Briefly, after fixation, slides were incubated with primary antibody at 37°C for 1hr and Discovery multimer detection system (Discovery OmniMap HRP, Discovery DAB, Roche) was used to detect and amplify immuno-signals. Primary antibodies: Ki67 (Thermo Fisher 14-5698-82), 1:500 dilution; Pan-CK (Novus Bio NBP1-48348H), 1:100 dilution; TREM2 (Proteintech 13483-1-AP), 1:150 dilution.

#### Tumor *in situ* hybridization

Staining was performed using the RNAscope platform (ACD), according to the manufacturer’s protocol for the RNAscope 2.5 HD Detection Reagent (red, 322360) and technical note for fresh-frozen tissue (320536). Tissue sections were fixed with 4% PFA for 15 mins at 4°C, and dehydrated with a series of ethanol washes (50%, 70%, 100%, 100%) for 5 minutes each. The sections were pretreated with hydrogen peroxide for 10 min, washed once with distilled water, pretreated with Protease IV for 30 min at RT and washed with 1X PBS. Sections were then individually hybridized for two hours at 40°C, with probes targeting either TREM2 (404111, ACD), DapB (negative control; 310043, ACD) or PPIB (positive control; 313911, ACD). After hybridization, sections were washed twice with 1X PBS for 2 minutes and subject to 6 amplification steps (30 min at 40°C, 15 min at 40°C, 30 min at 40°C, 15 min at 40°C, 30 min RT, 15 min RT) prior to detection. Signal was detected using Fast Red reagent (322360, ACD) for 10 minutes at RT, and briefly washing with tap water prior to counterstaining with hematoxylin. Slides were mounted using xylene and EcoMount. Images were scanned using a Leica-Aperio Versa slide scanner.

### Quantification and statistical analysis

#### Cohort genomic data quality control (QC)

##### UK Biobank (UKB)

UKB-provided measured genotype, imputed genotype (GRCh37, imputed data release 3) and phenotype data^1^ was accessed as part of application 58510. We selected subjects with available imputed genomic data (field 22028) and at least one paired creatinine (field 30700) and CyC measurement (field 30720), and excluded subjects with sex chromosome aneuploidy (field 22019), discordant genetic sex (fields 31 and 22001), excess heterozygosity and missing rate (field 22027). To classify genetic ancestry, we lifted over directly genotyped and linkage disequilibrium (LD)-pruned high-quality variants (bi-allelic SNPs, MAF > 0.1%, call rate > 99%) to GRCh38 and merged with variants available from an integrated callset (call rate > 95%) derived from 1000 Genomes and Human Genome Diversity Project (HGDP, gnomAD). LD pruning was implemented using PLINK1.9 with parameters ‘--indep-pairwise 50 5 0.2’. Principal components (1-10) were computed using the unrelated reference subjects (PC-relate kinship coefficient < 0.05) then projected onto all reference and UKB subjects. Next, a random forest classifier was trained using ancestry data from the reference cohort, implemented in the gnomAD package for Hail. This classifier was applied to the UKB subjects, and genetic ancestry was assigned with a minimum probability of 70% (Table S1). Relatedness data was extracted from the UKB-provided kinship matrix, generated using KING software. For the EUR ancestry group, subjects were split into a discovery cohort (n=381,764 subjects) and validation cohort (n=50,000 subjects), with the validation cohort comprising a random selection from unrelated UKB subjects (KING kinship coefficient < 0.0442). For all other ancestry groups, all subjects were used as discovery cohort. For all analyses using imputed data, we filtered to variants with INFO score > 0.8 and MAF > 1% across whole cohort.

##### Genotype-Tissue Expression (GTEx) project

Whole-genome sequencing data (GRCh38) and controlled-access metadata (including time of death) was accessed through dbGaP (phs000424.v8.p2) as part of application 26811. The provided imputed data had already undergone extensive quality control, however, we removed an additional 9 subjects with a PC-relate kinship coefficient > 0.05. We identified EUR ancestry subjects (n=678) as above using 1000G/HGDP reference data to train a random forest classifier that was applied to GTEX subjects, using high-quality LD-pruned common variants (bi-allelic SNPs, MAF > 0.1%, call rate > 99%, r^2^<0.1), LD pruning was implemented using the ‘ld_prune’ function in Hail, subsequent to removal of high-LD regions^100^. In this smaller cohort, ancestry was defined using a minimum probability of 50% followed by removal of PCA outliers with a PCA Z-score > 5.

##### The Cancer Genome Atlas (TCGA)

Germline array data (Birdseed format, GRCh37) was downloaded from the GDC Legacy archive as part of dbGaP application 26811, before conversion to VCF format. For sample QC, we started with a sample list defined by Sayaman et al^101^, which selected one germline sample per subject, prioritizing blood-derived or high call-rate samples, while removing samples with excess heterozygosity or hematological malignancies. For additional sample QC, we removed samples with discordant sex (using the impute_sex function in Hail), excess hetero- or homo-zygosity (Z-score > 3, using agg.inbreeding function in Hail), related subjects (PC-relate kinship coefficient > 0.05) and called genetic ancestry as described for the GTEX cohort (n=7260 EUR patients). For imputation in the unrelated EUR population, we selected variants with call rate > 95% and MAF > 0.1%. Imputation was performed using the TOPMED server, which automatically lifts over variants to GRCh38. For the final cleaned dataset, we selected autosomal variants imputed with r^2^ > 0.6 and MAF > 0.1%.

##### Stockholm Tartu Atherosclerosis Reverse Networks Engineering Task (STARNET)

STARNET is a cohort of 600 Caucasian patents of Eastern European origin, with a confirmed diagnosis of coronary artery disease. Genomic data quality control has been described previously^51^. Briefly, array-based genotyping was performed on germline DNA from blood, followed by imputation against the 1000 Genomes phase 1 SNPs. Comparison of population structure with 1000 Genomes cohort confirmed that all STARNET subjects had European genetic ancestry.

##### Immunotherapy meta-cohort (panIO)

We requested access to 8 cohorts of patients treated with CPI (anti-PD-1, anti-PD-L1 and/or anti- CTLA4) with available germline exome sequencing and clinical outcome (Table S2a-b). Clinical annotations were downloaded from the supplemental data from associated manuscripts or requested directly from principal investigators. Samples were excluded if there was insufficient data to report at least one outcome measure (overall survival, progression-free survival, durable clinical benefit). Durable clinical benefit (binary) was defined by patients with no radiological progression > 6 months or overall survival > 1 year. Harmonized germline short variant calling was implemented using nf-core/sarek pipeline, with Strelka mutation caller^102^ and GRCh38 reference genome. gVCFs were merged using Illumina gvcfgenotyper tool and imported into Hail for processing. Samples with discordant sex (n=13) were identified by comparison of sex reported in clinical metadata and genetic sex determined from integration of X chromosome heterozygosity and Y chromosome genotype counts (via PLINK 1.9 impute-sex function). For the small minority of patients without supplied sex (n=4), sex was genetically imputed. For variant QC, calls filtered by Strelka were removed, SNPs calls required a minimum depth of 7 while indel calls required a minimum depth of 10. Each variant required a call rate >90% and at least one ‘high-quality’ call defined as one homozygous ALT call or one heterozygous ALT call (with allele balance >15% for SNP or >20% for indel). Samples with a call rate <90% or excess hetero- or homozygosity (Z score > 3) were removed. No subjects had >3^rd^ degree relatedness, which also excludes the possibility of duplicates samples in the cohort. EUR ancestry subjects were identified as for GTEX cohort. Imputation of EUR population was performed using the TOPMED server. For the final cleaned dataset, we selected autosomal variants imputed with r^2^ > 0.6 and MAF > 0.1%.

#### Computation of principal components

We computed 20 principal components (PCs) on all subjects (including related) and all genotyped variants, as per the BOLT-LMM manual, implemented in PLINK2 (--pca function). Due to computational complexity, the PLINK2 PCA approximation (--approx) was used for the EUR population. To account for genetic ancestry in downstream analyses, PCs (1-4) were computed on high-quality linkage disequilibrium (LD)-pruned variants (bi-allelic SNPs, MAF > 0.1%, call rate > 99%, r^2^<0.1), with SNPs in known high-LD regions removed^100^. For UKB, high-quality SNPs were derived from the ‘in_PCA’ field from the UKB-provided SNP QC file. In the UKB cohort, PCs were computed with related subjects removed (approach described above), and then projected onto all remaining samples, using the ‘run_pca_with_relateds’ function in the gnomAD package for Hail. In other cohorts (where related subjects were removed), PCs were computed using the ‘hwe_normalized_pca’ function in Hail.

#### Genome-wide association analysis (GWAS)

eGFR-CyC and eGFR-Cr were calculated using CKD-EPI equations^54^ implemented in the nephro package for R, with race term set to 0 for all subjects. For subjects we more than 1 paired creatinine and CyC measurement, we selected the earliest complete datapoint. Genome-wide association analyses (GWAS) in the discovery cohorts (for eGFR-CyC and eGFR-Cr were performed in each ancestry group, including related subjects, using BOLT-LMM^8^ with covariates including age (field 21003), age^2^, sex (field 31), genotyping array (binarized from field 22000), recruitment centre (field 54), and genetic PCs 1-20 (described above). LD score matrices for each ancestry group were downloaded from the Pan-UK Biobank project (https://pan.ukbb.broadinstitute.org/). To assess for confounding we determined the attenuation ratio of each trait via LD score regression, which was within the expected range for polygenic traits (Table S3)^103^.

#### Structural equation modelling

Structural equation modelling of eGFR-Cr and eGFR-Cy summary statistics was implemented in the Genomic-SEM package for R^33^ and performed as per the GWAS-by-subtraction tutorial (https://rpubs.com/MichelNivard/565885). Briefly, for EUR, AFR and CSA populations, we performed LD score regression using LD matrices from the Pan-UK Biobank Project (https://pan.ukbb.broadinstitute.org/). We designed a structural equation model (summarized in Figure 1c), with latent traits estimated using the userGWAS function parallelized across each chromosome. Summary statistics for each latent trait (renal function, CyC-production) were extracted and effective sample sizes were estimated using the script provided by the Genomic- SEM authors (https://github.com/GenomicSEM/GenomicSEM/wiki/5.-User-Specified-Models-with-SNP-Effects). CyC-production summary statistics were standardized by setting A1 as the GRCh37 ALT allele and A2 as the GRCh37 REF allele, and multiplying the effect size of CyC- production by -1 so a higher effect size reflects increased CyC production.

#### Processing of summary statistics

Clumping was performed in the EUR eGFR-CyC summary statistics, implemented in PLINK 1.9 with parameters clump-r2 0.001, clump-p1 5e-8, clump-p2 5e-8 and clump-kb 10000 using 1000 Genomes reference data (derived from European subjects). For each clump, the index SNP (SNP with lowest p value) was annotated using the OpenTargets Genetics (https://genetics.opentargets.org/) variant-to-gene pipeline^104^, which integrates both proximity and functional genomics data. For the small minority of variants (n=2) not represented in the OpenTargets database, the index SNP was annotated to the nearest coding gene. Partitioned heritability analysis was performed using the LDSC package for R^105^ using the provided datasets, as per the tutorial by the package authors (https://github.com/bulik/ldsc/wiki/Cell-type-specific-analyses). For each trait and tissue-sample pair, we extracted the t-statistic as the ratio of the coefficient and standard error. To compare cell type-specific enrichment between renal function and CyC-production latent traits, we computed the absolute difference in t-statistic between eGFR-CyC and each latent trait, for each tissue sample. Colocalization analysis was performed using the coloc package for R^50^, using the single-variant assumption. Gene set enrichment analysis of CyC-production latent trait was performed using MAGMA^106^, implemented in the FUMA web server (https://fuma.ctglab.nl/) with a 0kb gene window. Mendelian randomization analysis, using cis-eQTLs probes for *CST3*, was implemented in GCTA-SMR^107^ using SMR- formatted eQTL data from the eQTLGen Consortium^58^.

#### Derivation and application of polygenic scores

CyC-production polygenic scores (PGS) were derived using LDpred2^44^ (automatic model) according to the package vignette (https://privefl.github.io/bigsnpr/articles/LDpred2.html). For the genome-wide score, HapMap3 variants were intersected with high-quality genomic variants available for all of UKB (array), TCGA (array) and GTEX (WGS) cohorts (n=1,031,527). For the exome-wide score, HapMap3 variants were intersected with high-quality exonic variants from the panIO cohort (n=352,549). The provided UKB LD reference was used for PGS derivation. Model fitting was confirmed by visual inspection of chain convergence for each PGS. The PLINK2 linear scoring function (--score) was used to apply the PGS to each cohort and to avoid exclusion of duplicate dbSNP IDs, the source data was filtered to the PGS variants according to position and alleles. The sample-level PGS was normalized by Z-scoring in each cohort. To generate a patient-level surrogate for CyC production, we modelled eGFR-CyC as a function of eGFR-Cr and sex, with intercept set as 0. We computed the residual of this model, termed CyC-residual, which is multiplied by -1 so that increasing CyC-residual reflects increased serum CyC relative to creatinine.

#### Functional genomics

ChIP-seq (for GR/NR3C1, timeseries accession: ENCSR210PYP) and ATAC-seq (timeseries accession: ENCSR385LRX) data for A549 cells treated with dexamethasone was downloaded from the ENCODE data portal (https://www.encodeproject.org/). Data was processed using the ENCODE data analysis pipeline, generating a p-value for each signal peak that reflects enrichment of DNA sequences. Data at the *CST3* locus was plotted using the karyoploteR package^108^ for R. Enhancer activity scores (‘ABC scores’) derived from the validated activity-by-contact model^93^, applied to 131 biosamples, was downloaded from ftp://ftp.broadinstitute.org/outgoing/lincRNA/ABC/Nasser2021-Full-ABC-Output/. Scores for the distal enhancer element at the *CST3* locus reflecting analysis of data derived THP-1 cells were extracted, and data from THP-1 cells treated with PMA was compared to data from naïve THP-1 cells.

#### Gene expression profiling

For GTEX and ENCODE gene expression profiling, gene-level counts derived from STAR-aligned RNA sequencing (RNA-seq) reads were downloaded from the GTEX (https://GTExportal.org/home/datasets) and ENCODE (timeseries accession: ENCSR897XFT) data portals respectively. TMM and library size normalization were applied using the edgeR package^109^ for R, generating TMM-normalized log-counts per million (CPM) expression values that can be compared between samples. For TCGA gene expression profiling, batch- and expression quantile-normalized data (RNA-seq) was downloaded from the PanCancer Atlas repository (https://gdc.cancer.gov/about-data/publications/pancanatlas). For STARNET gene expression profiling was performed as previously described^108^ – briefly, gene counts were adjusted for GC content, library size and quantile-normalized implemented in EDAseq^110^, prior to log-transformation. For digital cytometry analysis implemented in CIBERSORTx^67^ (for STARNET and GTEX cohorts), gene expression was normalized to gene length to generate transcripts per million (TPM) expression values. CIBERSORTx was run in absolute mode with LM22 reference set, 100 permutations and B-mode batch correction.

#### Expression quantitative loci (eQTL) analysis

eQTLs were identified in the STARNET^51^ cohort using the Kruskal Wallis test statistic (additive model), as implemented by the tool kruX^111^, using individual-level genotype and gene expression data (data processing described above). To identify associations between *CST3* and SNPs present at the *SERPINA6/ SERPINA1* loci, we carried out this association analysis using 72 SNPs previously shown to be significantly associated with plasma cortisol^49^. This approach was applied to all non-vascular tissues (n=5) in the STARNET cohort (subcutaneous fat, visceral abdominal fat, skeletal muscle, liver, blood). As the 72 SNPs reflecting 4 independent LD blocks, we modelled this analysis as 20 (4 LD blocks, 5 tissues) independent hypotheses and so the Bonferroni-corrected significance threshold was 0.0025. For independent validation of the significant eQTL associations in GTEX, we performed Kruskal Wallis tests in two tissues (visceral adipose fat, liver) using the ‘kruskal.test’ function for R, using individual-level genotype and gene expression data (data processing described above). For further characterization of significant eQTLs, we constructed a recessive linear model of *CST3* gene expression as a function of genotype (binarized to 0/1 versus 2), using the ‘lm’ function for R.

#### Single-cell RNA sequencing (scRNA-seq) analysis

For analysis of scRNA-seq profiles of human skin tumors, scRNA-seq expression matrices and metadata for Jerby-Anon et al.^97^ and Yost et al.^77^ were downloaded from Single Cell Portal (accession SCP109) and GEO (accession GSE123813), respectively. For analysis of scRNA-seq profiles of spleen and PBMCs, scRNA-seq expression matrices were downloaded from For Yost et al. the peritumoral T cell-specific samples were excluded from the analysis. Count or normalized expression data was imported into Seurat^112^ (version 4.0), filtered (according to number of features, <10,000, and mitochondrial content, <7.5%, per cell), log-normalized (if applicable) and scaled. Highly variable features (n=2000) were used for principal component analysis followed by clustering (Louvain algorithm). Immune clusters were annotated by comparison to reference PBMC data, implemented in clustifyR package for R^91^. Unannotated clusters (presumed to reflect one of tumor, cancer-associated fibroblast or endothelial cells) were manually annotated via established marker gene expression^98^ and clonal copy number variation profiles, examined using the inferCNV^113^ package for R. Patient-level pseudobulk cluster-specific expression data was extracted using the ‘AverageExpression’ function in Seurat.

For analysis of scRNA-seq profiles of murine Mm1 tumors, cellranger-processed data was imported into Seurat^112^ (version 4.0). Quality control steps included removal of putative doublet cells (implemented in DoubletFinder^114^) and removal of cells with >20% mitochondrial genome-aligned reads or fewer than 200 features (UMIs). Data processing in Seurat included normalization, scaling (regressing out the effect of cell cycle genes), integration, Louvain clustering, dimensionality reduction and visualization. Each cluster was annotated as one of 14 cellular populations according to expression of validated marker genes (summarized in Table S7, and Figure S7a) Differential gene expression was identified from pseudobulk data, implemented in the Libra package for R^115^. Differentially enriched/depleted cell populations were identified by modeling logit-transformed cell proportions as a function of tumor genotype, implemented in the speckle package for R^71^.

To assess changes in the monocyte population following dexamethasone, we reanalyzed whole blood scRNA-Seq datasets from ICU-admitted COVID-19 patients treated with or without dexamethasone^64^. Initial data pre-processing and cell identity annotations were performed as described previously. Briefly, cell identity annotations were generated by mapping single-cell transcriptomes to the PBMC scRNA/CITE-seq multi-omic reference (Azimuth)^112^. To identify a high-confidence TREM2+ monocyte population, we extracted cells annotated as CD14+ monocytes and repeated batch effect correction (implemented using the ‘FindIntegrationAnchors’ function in Seurat), dimensionality reduction, and clustering. High-confidence Trem2+ monocytes were identified by expression of known TREM2+ monocyte markers (TREM2, APOE, CSF1R^76, 85^), identified by gene-weighted density estimation implemented in the Nebulosa package for R^73^. Differentially enriched/depleted cell populations were identified as above.

#### Cosinor regression

Cosinor regression for blood markers (bilirubin, CyC-residual) and gene expression (*FKBP5*, *CST3*) as a function of time was performed using the cosinor package for R. A cosinor model has 4 parameters – MESOR (intercept), period (assumed as 24 hours), amplitude and acrophase (timing of activity peak). Bilirubin was extracted from UKB field 30840 and CyC-residual was derived as above; both metrics were standardized with Z-scoring stratified by age (decade) and sex. Gene expression was derived from TMM-normalized TPM data (implemented in edgeR), to facilitate intra- and inter-sample comparisons. For UKB, time referred to time of sampling and for GTEX, time referred to time of death; both rounded to the nearest hour in 24-hour clock. Amplitude coefficients were extracted from the transformed coefficients table.

#### UK Biobank cancer cohort

To identify patients who were treated with non-topical exogenous GCs, we reviewed field 20003 for coded medications bio-equivalent to dexamethasone or prednisolone. Subject lifespan was extracted from analysis of fields 40007 and 34. Parental lifespan was extracted from analysis of fields 2946, 1845, 1797, 1835, 1807 and 3526. Using cancer registry data (fields 40005, 40012, 40008, 40011), ICD10-coded cancer diagnoses were extracted and mapped to Phecodes (https://phewascatalog.org/). Using a curated list of operation codes (OPCS-4) reflecting curative procedures for 13 main tumor groups (Table S4), we mapped each cancer diagnosis to matched surgeries that occurred no more than 90 days prior to the coding entry. To account for variation in operation data availability prior to 2000, we filtered the data to cancers that were diagnosed after the year 2000. In cases where a patient was coded with a cancer of the same primary type more than once, the entries were merged. Patients with more than one discrete cancer diagnosis were excluded (n=2435 subjects) due to the difficulty in defining the time since diagnosis. For recruited patients who had died, we manually reviewed details from the death certificate (field 40010) to identify descriptions that were consistent with cancer-specific mortality.

#### Survival analyses

For Cox regression of overall and cancer-specific survival against CyC-residual, the time variable used was time from blood sampling to death or last follow-up date (nominally June 2020). For subjects with multiple CyC-residual datapoints over time, each datapoint was annotated with survival time relative to blood sampling and treated independently. Model covariates included age (at blood sampling), sex, body mass index (BMI), hemoglobin, eGFR-Cr, C-reactive protein. For Cox regression of lifespan (for subject and parents) against CyC-production PGS in the UKB validation cohort, we used age at death or age at most recent follow-up as the time variable. Model covariates included year of birth (of subject, as parental birth years are not recorded) to account for historical increases in mean lifespan.

For Cox regression of cancer-specific survival against CyC-production PGS, it was necessary to consider bias from left truncation, where patients who died between diagnosis and the recruitment period would not be recruited. To account for this, the time interval used for Cox regression of overall and cancer-specific survival against CyC-production PGS in UKB referred to time from recruitment to death or data cut-off (June 2020). In contrast, TCGA patients were generally recruited close to the time of cancer diagnosis, prior to surgical resection of tumor and so time from diagnosis to death or last-follow-up date was used. Cancer-specific survival was extracted from the ‘DSS’ and ‘DSS.time’ fields in the TCGA clinical data resource available as part of the PanCancer Atlas (https://gdc.cancer.gov/about-data/publications/pancanatlas). Cancer- specific survival analyses with respect to CyC-production PGS were adjusted for age of diagnosis, genetic ancestry (PC1-4), sex (except sex-specific cancers), and a term reflecting whether curative surgery was performed. For UKB this term was derived from matching with curative operation codes as described above, for TCGA this term was derived from the field ‘residual_tumor’ in the clinical data resource. UKB-specific PGS-cancer survival analyses were additionally adjusted for recruitment center (to account for regional heterogeneity in cancer outcomes) and genotyping array. Pan-cancer inverse variance-weighted meta-analysis in each cohort (UKB, TCGA) was implemented in the meta package for R^92^ using both fixed and random effects models.

For phenome-wide time-to-event analysis in UKB, all UKB ‘first occurrence’ fields and cancer registry data (fields 40005 and 40006) were extracted, with ICD10 codes mapped to Phecodes. If multiple ICD10 codes mapped to a single Phecode, the earliest date of diagnosis was selected. For each time-to-event Phecode, the time variable was defined as time from birth to first occurrence of diagnosis or most recent follow-up date. To account for region-specific variability in health record linkage, this date was determined by either the most recent coded diagnosis or most recent UKB center visit. Each phenotype-specific Cox regression was adjusted for sex, genetic ancestry (PC1-4) and year of birth (to account for historical variation in disease risk).

#### Digital pathology analyses

Digital image analysis of H+E stains as well as Trem2, panCK and Ki67 IHC were performed using HALO™ digital image analysis software version v3.4.2986.151 (Indica Labs, Corrales, NM, USA). All H&E scans were reviewed by two pathologists (DL and VHK), and the regions of viable respectively necrotic tissue was determined and computed. Necrotic tissue was excluded for further analysis. In order to analyze the tumor/stroma interaction, a deep neural network algorithm was trained on the panCK scans to recognize epithelial and non-epithelial compartments. Graphical overlays for both compartments simplified the quality control of the tissue classifier. The total area of the two compartments was then calculated automatically.

The ‘Multiplex IHC v3.1.4’ algorithm of HALO™ was implemented for analysis of Ki67 and Trem2 IHC. The nuclei and the chromogens were detected by color deconvolution with thresholds determined by internal controls. For Ki67 IHC, a proliferation index (Ki67+ cells / total cells) was calculated. For Trem2 IHC, the proportion of Trem2+ per unit tissue area was calculated, and the distance of each Trem2+ cell from the tumor border was measured using the proximity analysis tool of HALO™.

#### Statistical analysis

Significance testing refers to two-tailed unpaired t-tests with the assumption of unequal variance unless stated otherwise. For bi-flank tumor experiments, differences were assessed using two-tailed paired t-tests. For statistical and computational analyses, we used R (version 4.0.2) and Python (version 3.7.4) implemented as a Jupyter Notebook.

